# Neuromotor Modules Revealed by Intraoperative Direct Electrical Stimulation of the Human Primary Motor Cortex

**DOI:** 10.1101/2025.10.20.25338233

**Authors:** Jodie J. Xie, Subing Huang, Kelvin Y. S. Lau, Amy H. S. Kong, Rosa H. M. Chan, Peter Y. M. Woo, Vincent C. K. Cheung

## Abstract

Muscle synergies extracted from multi-muscle electromyographic (EMG) signals are widely interpreted as fundamental building blocks of motor control which coordinate groups of muscles during movement. Whether EMG-derived muscle synergies represent genuine neurally encoded modules or motor regularities arising from task or biomechanical constraints has been hotly debated, because neurophysiological data that definitively demonstrate the neural basis of muscle synergies in humans has been lacking. Here, we seek to validate the potential neural origin of upper limb muscle synergies by exploiting direct electrical stimulation (DES) of the primary motor cortex (M1) routinely delivered by neurosurgeons during awake craniotomy surgery for glioma excision. Across 13 patients, 69% of the muscle synergies observed during pre-operative voluntary behaviors could be matched to DES-evoked muscle synergies or their combination. Analysis of the synergies’ cortical activity maps further revealed that the cortical representations of the sparser muscle synergies were more anterior and distributed, and those of the non-sparse synergies, more posterior and localized. Our results not only provide direct causal evidence arguing for the neural origin of most behavioral muscle synergies in humans, but also demonstrate the existence of two M1 subdivisions with distinct patterns of muscle synergy organization.

## INTRODUCTION

Humans are endowed with the extraordinary ability to execute a wide range of movement, which requires a highly coordinated central nervous system (CNS) to translate any motor intention to muscle activities. Understanding how the CNS handles the complexity of muscle coordination for diverse voluntary behaviors has remained a central, hard question in neuroscience (Bernstein, 1967; Bizzi and Ajemian, 2015). When performing motor tasks, the CNS may address this complexity by generating muscle patterns through the combination of a small number of pre-organized, low-level motor modules (Bizzi et al., 1991; Tresch et al., 1999b; Hart and Giszter, 2004), which may be identified as muscle synergies through electromyographic (EMG) signal decomposition. These motor modules may serve as the fundamental building blocks of movement where muscle groups are co-activated together (d’Avella et al., 2003; Roh et al., 2011; Saltiel et al., 2001; Ting and Macpherson, 2005; Tresch et al., 2002).

It has been argued that algorithm-decomposed muscle synergies may not correspond to the actual motor modules encoded in the CNS (Cheung and Seki, 2021; Ting et al., 2015; Tresch and Jarc, 2009). Muscle synergies extracted from EMGs may represent constraints from experimental tasks, biomechanical constraints arising from neuro-musculoskeletal functions and anatomy, or simply epiphenomena of other motor-control principles (Alessandro et al., 2020; Berniker et al., 2009; De Groote et al., 2014; McKay and Ting, 2012; Kutch and Valero-Cuevas, 2012; Todorov and Jordan, 2002). To resolve these controversies, numerous studies have relied on neuronal recordings in animal models, including the frog (Hart and Giszter, 2010), mouse (He et al., 2025), and monkey (Takei et al., 2017), to demonstrate the role of the spinal interneuronal network in the organization of muscle synergies, and to validate their neural origin. Specifically, spike-triggered averaging of EMGs have been successfully applied to spinal unit recordings to estimate EMG-derived behavioral muscle synergies, directly demonstrating their neurophysiological basis (Takei et al., 2017; He et al., 2025). Other animal studies have relied on focal electrical or optogenetic stimulations, applied to the spinal cord of the frog (Bizzi et al., 1991; Saltiel et al., 2016), mouse (Salmas and Cheung, 2023; He et al., 2025), as well as the motor cortex of the cat (Ethier et al., 2006; Capaday, 2022) and monkey (Holdefer and Miller, 2002; Huffmaster et al., 2017; Overduin et al. 2012), to independently retrieve behavioral muscle synergies. Results from these studies have shown that their neural representations may have at least a coarse topographical organization.

To what extent the muscle synergies observed in human motor behaviors are indeed organized by the CNS is still a contentious topic in motor neuroscience. Early indirect evidence supporting the neural origin of muscle synergies in humans was derived from the observation that muscle synergies remain stable after the task biomechanical requirements change (Torres-Oviedo and Ting, 2010), and during motor adaptation when limb perturbations are compensated by recruiting the same synergies (Berger et al., 2013). More recently, non-invasive brain mapping tools such as functional magnetic resonance imaging (fMRI), electroencephalography (EEG), and transcranial magnetic stimulation (TMS) have shown the presence of cortical “synergy access points” (Rana et al., 2015) that provide access to coordinated muscle synergies apparently encoded in the human CNS. Muscle coordination patterns can be inferred from distributed cortical representations obtained by fMRI and TMS (Yani et al., 2018) or from EEG-to-EMG corticomuscular coherence (Witham et al., 2011; Zandvoort et al., 2019), or can be activated by focal TMS applied to specific motor cortical loci (Gentner and Classen, 2006; Fricke et al., 2020; Yarossi et al., 2022). Nonetheless, results of these non-invasive experiments are spatially and/or temporally imprecise when compared with those from recordings or stimulations of individual neurons. Several recent human studies have argued against the neural basis of muscle synergies, showing that a fixed set of low-dimensional basis vectors derived from factorization of EMGs from a few behavioral tasks are insufficient to explain the whole gamut of variable movement in the task space (Barradas et al., 2020; De Rugy et al., 2013; Zelik et al., 2014). According to this view, muscle synergies could be byproducts of non-neural factors or choices of analytic methods rather than from neuromotor constraints that can account for motor variability (Inouye and Valero-Cuevas, 2016; Steele et al., 2015).

An alternative way of studying the human motor system is by direct electrical stimulation (DES) of the motor cortex, a technique routinely delivered by neurosurgeons during awake craniotomy brain surgery for glioma resection. Not only does this procedure serves as a gold standard for brain mapping and for preserving essential function during tumor resection, but it also presents a window of opportunity to address fundamental scientific questions in neuroscience with both higher spatial and temporal resolution than non-invasive approaches (Borchers et al., 2012; Fornia et al., 2020a; Rech et al., 2019; Southwell et al., 2016; Servick, 2022; Viganò et al., 2019). During an awake craniotomy, CNS loci yielding positive functional responses following DES indicate the regions of interest that must be spared for functional preservation. For motor cortical stimulation, the motor functional and neurophysiological responses elicited by DES should at least indicate the muscle patterns accessible from this stimulated locus, thereby providing insights on its attendant functions. This approach of exploiting DES to study the motor system goes back to the pioneering works of Fritsch and Hitzig (1870) (Gross, 2007), John Hughlings Jackson (Meares, 1999), and Wilder Penfield (1937). However, it remains viable and highly relevant today as new tools for more precise stimulation, recordings, and large-scale data analytic methods become available. Recent advances in both invasive and non-invasive brain mapping techniques have refined our understanding of the somatotopic representations of the human primary motor cortex (M1), somatosensory cortex, and cerebellum (Catani, 2017; Fan et al., 2016; Gordon et al., 2023; Mottolese et al., 2013; Petersen et al., 2024; Roux et al., 2018; Roux et al., 2020).

In this study, we aim to obtain neurophysiological evidence for the existence of muscle synergies in humans. We asked whether upper limb muscle synergies observed in daily activities can be independently evoked by applying DES to the human M1 during a standard awake craniotomy procedure for glioma resection. This approach offers an invaluable opportunity to study the human motor system without incurring any additional risk to the patient. We evaluated the similarity between muscle synergies elicited by DES and behavioral muscle synergies extracted from EMGs recorded during pre- and post-operative upper limb voluntary motor tasks. We also mapped the motor cortical loci whose stimulations would activate the same synergy to gain insights into the synergies’ topographical organizations in M1. Our findings may not only be the most direct casual demonstration to date of the neural origin of muscle synergies in humans, but also reveal distinct M1 subdivisions with different synergy organizational patterns, shedding light on the long-standing controversy on how M1 may be organized in relation to different muscle synergies.

## RESULTS

Patients diagnosed with gliomas with no clinically apparent motor functional impairment were recruited (n = 13). All subjects underwent, as part of their treatment plan, an awake tumor resection surgery that necessitated the exposure of at least part of the upper limb region of the M1. They also participated in three data recording sessions (Figure 1A). In the pre- and post-operative (pre- and post-op) sessions, surface electromyographic signals (EMGs; 12-15 muscles of contralateral arm) were recorded during 10-11 upper limb tasks related to activities of daily living. In the intraoperative session after the brain was exposed, the patient was awakened and asked to stay at rest or perform specific motor tasks as direct electrical stimulation (DES; 2-5 mA, 60 Hz, 1-3 s per trial) was sequentially delivered to multiple cortical loci. This motor mapping procedure, performed under neuro-navigation, permitted the identification of the loci whose stimulations would elicit involuntary muscle responses (“positive” loci), and would be preserved during tumor resection. Surface upper limb EMGs were collected during intraoperative motor mapping. Non-negative matrix factorization (NMF) was then conducted to extract behavioral muscle synergies from the pre- and post-op EMGs, and DES-evoked muscle synergies from the intraoperative EMGs elicited from positive stimulation loci (Fricke et al., 2020; Huffmaster et al., 2017; Yarossi et al., 2022) (Figure 1B). For each DES-evoked synergy, the variation of the synergy’s time- and trial-averaged coefficient across the stimulation loci further allowed us to construct a cortical map of the synergy’s activities (see below).

**Figure 1.**
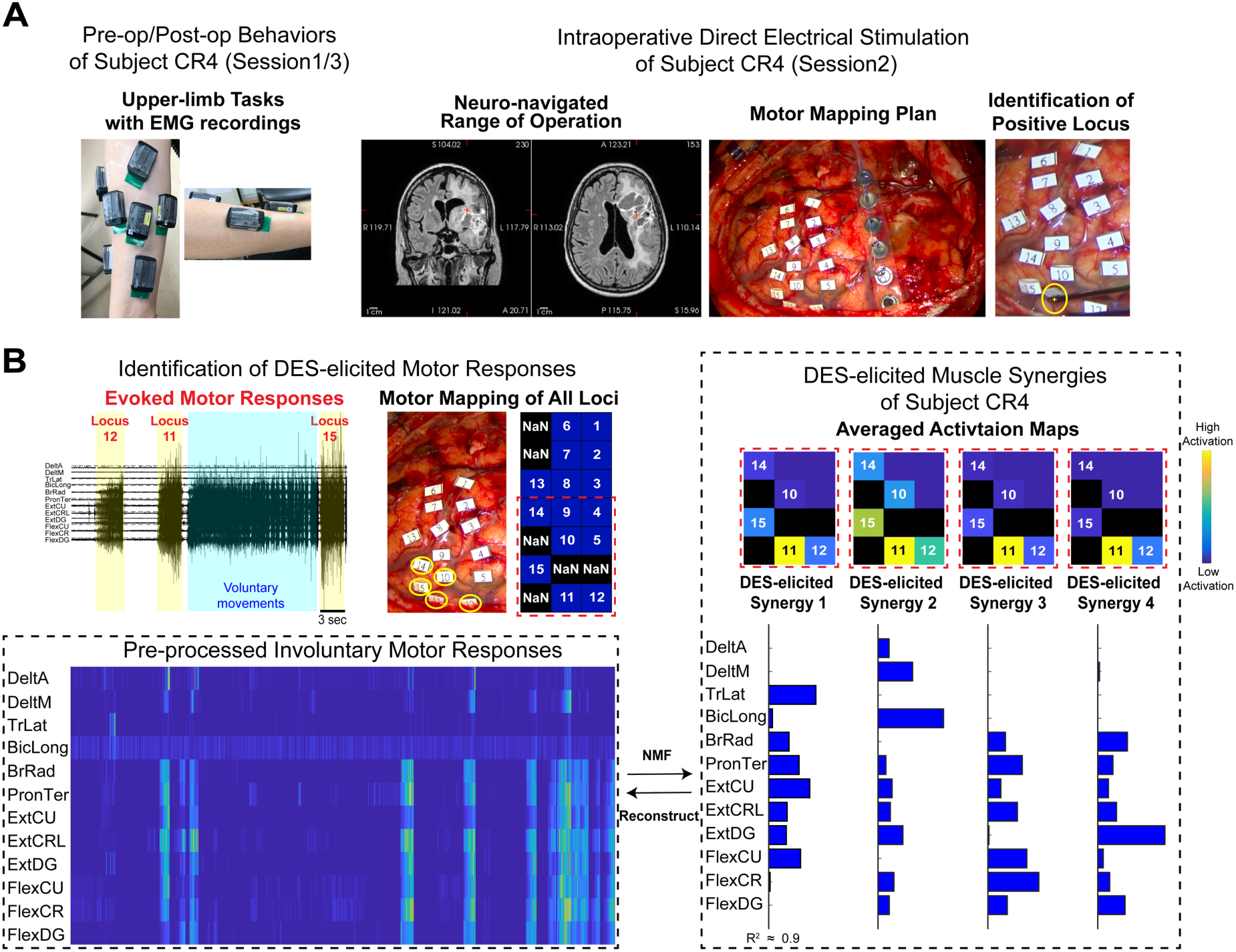
Experimental sessions and preprocessing of the muscle patterns evoked by direct electrical stimulation (DES) of the human primary motor cortex (M1). **(A)** In the pre-operative (pre-op, session 1) and post-operative (post-op, session 3) behavioral sessions, surface electromyographic signals (EMGs; 12-15 contralateral muscles) were recorded from 10-11 upper limb tasks related to daily activities. In the intraoperative (session 2) session, motor mapping, a standardized neurosurgical procedure performed in glioma patients undergoing awake craniotomy surgery, was performed, so that the cortical tissues surrounding the loci that yielded positive motor responses upon stimulation (yellow + in rightmost panel) would be spared from excision for the preservation of motor functions post-op. The EMGs collected from session 2 were elicited from M1 loci with bipolar DES (2-5 mA, 60 Hz, 1-3 s) while the subject was at rest but awake, or performing motor tasks. **(B)** The DES-evoked muscle patterns (from loci 12, 11, and 15 in this example) were first parsed out from the continuously recorded EMG trace (upper left panel). In this example from subject CR4, out of the 15 labelled loci, positive motor responses could be evoked from 5 positive loci (loci 10, 11, 12, 14, 15, highlighted by yellow circles and a rectangle with red dashed borders). All DES-evoked EMGs were then aggregated into a single data matrix and preprocessed (lower left panel). The non-negative matrix factorization (NMF) algorithm was then applied to the preprocessed EMG to identify the muscle synergies. For CR4, the DES-evoked EMGs could be reconstructed by linearly combining 4 muscle synergy vectors (lower right panel), each scaled by activation coefficients whose magnitudes across the positive loci are depicted here as a heat map (upper right panel). In this heat map, each square denotes a stimulated cortical locus, and the relative positions of the squares match the relative anatomical locations of the loci on M1.

### Behavioral muscle synergies could be explained by DES-evoked synergies

We began our analysis with an evaluation of the robustness of the extracted behavioral muscle synergies by comparing the pre- and post-op synergies. Since in all subjects the tissues in the vicinity of the positive loci were spared during tumor resection (Bello et al., 2008; Penfield and Boldrey, 1937; Mansouri et al., 2023; Yingling et al., 1999), none of the subjects reported any noticeable post-op long term motor functional impairment. We therefore presumed the pre- and post-op muscle synergies to be similar. From all subjects, we identified 64 pre-op and 50 post-op muscle synergies, categorizable by *k*-means into 5 pre-op and 10 post-op synergy clusters, respectively (Figure 2A). All pre-op clusters could be matched to a post-op cluster with high scalar product (SP) between their cluster centroids (0.94 ± 0.07, mean ± SD; range of 0.81 to 0.98), with SP values higher than those from matching the centroids of clusters of shuffled pre- and post-op synergies (0.72 ± 0.11, mean ± SD; range of 0.60 to 0.89). When the pre- and post-op synergies of each subject were matched to each other, 20% of the post-op synergies (10 of 50) could not be well matched (SP < 0.65 ± 0.09, mean ± SD). These synergies also tended to be sparser vectors with more muscle components closer to zero, consistent with the observation that on average, the post-op synergies had slightly higher sparseness values than the pre-op synergies (pre-op cluster centroids, sparseness of 0.61 ± 0.14; post-op, 0.66 ± 0.18; mean ± SD).

**Figure 2.**
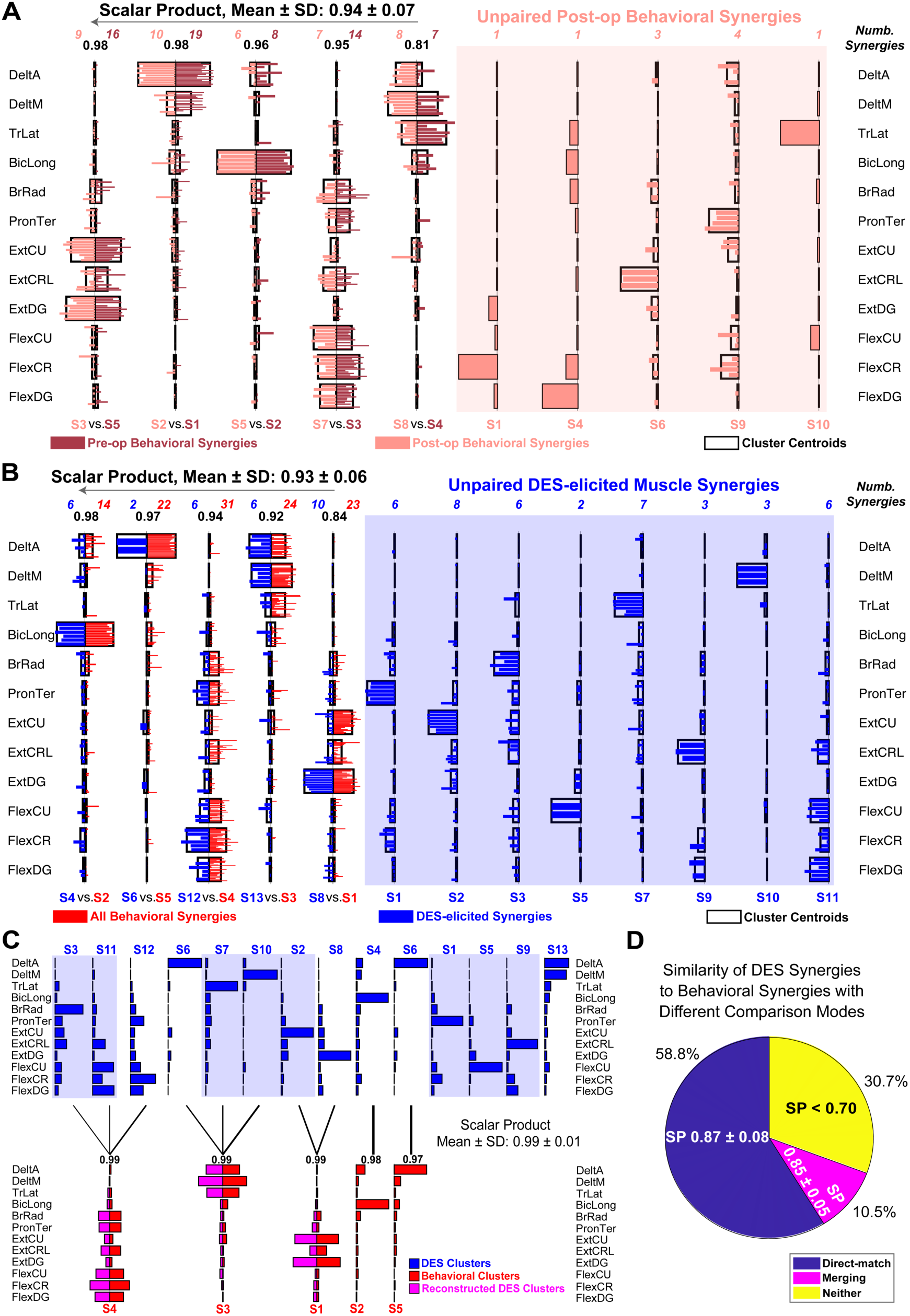
The behavioral and DES-evoked muscle synergy clusters were comparable. **(A)** We first *k*-means-clustered and compared the pre-op (dark pink) and post-op (light pink) muscle synergies. All 5 pre-op clusters could be matched to a post-op cluster with moderate to high scalar product similarity (0.94 ± 0.07, mean ± SD). Besides, 5 unpaired post-op clusters (synergies in light red background), each containing very few synergies (1 to 4), were observed. In each cluster, plots of individual synergy vectors (bars with filled colors) are superimposed onto the bars that denote the cluster centroid (bars without color). The scalar product similarity between each pair of matched cluster centroids is shown above the synergies in black; the numbers of pre- and post-op synergies classified into each cluster are shown above the synergies in dark red and light red, respectively. **(B)** The behavioral muscle synergy clusters (pre-op and post-op combined, red) could be matched to clusters of DES-evoked muscle synergies (blue) with scalar product of 0.93 ± 0.06 (mean ± SD). Besides, we observed 8 unpaired DES-specific clusters (synergies in blue background), most of which were dominated by the activity component of a single muscle. **(C)** Some of the behavioral synergy cluster centroids could be accounted for with higher similarity values when matched to vectors that were produced by linearly combining multiple paired and unpaired DES-evoked cluster centroids. For example, combining the DES-evoked clusters S3, S11 and S12 (blue) results in a merged vector (magenta) that could be matched to the behavioral cluster S4 (red) with a scalar product of 0.99. As in (B), unpaired DES clusters are plotted with blue background. **(D)** When the behavioral and DES-evoked synergies of each subject were compared, 59% of the behavioral synergies (including pre- and post-op synergies) could be directly matched to their corresponding DES-evoked synergies with scalar product ≥0.7, 10% could be accounted by for merging multiple DES-evoked synergies, and 31% could not be explained by either direct one-to-one matching or merging (scalar product <0.7).

To determine whether the behavioral muscle synergies could be independently retrieved by focal M1 stimulations, we evaluated how well the DES-evoked synergies could account for the pre- and post-op synergies. From all subjects, 71 DES-evoked synergies were identified and grouped into 13 clusters by *k*-means (Figure 2B). Given the generally high similarity between pre- and post-op muscle synergies (see above), we further identified 5 behavioral synergy clusters from the combined pre- and post-op synergy set (n = 114). The centroids of these 5 behavioral synergy clusters could be paired with those of 5 DES-evoked clusters with SP of 0.84 to 0.98 (0.93 ± 0.06, mean ± SD) (Figure 2B), which was higher than the baseline SP between cluster sets derived from shuffled DES-evoked and behavioral synergies (0.76 ± 0.16 , mean ± SD; range of 0.50 to 0.92). When the behavioral (pre-op and post-op sessions) and DES-evoked synergies of each subject were compared, 58% of the latter (41 of 71) could not be well matched to a corresponding behavioral synergy (SP of 0.69 ± 0.13, mean ± SD). These 41 synergies also appeared to be sparser than the behavioral synergies, consistent with the observation that on average, the DES-evoked synergies had higher sparseness (behavioral cluster centroids, sparseness of 0.58 ± 0.17; DES-evoked, 0.65 ± 0.15; mean ± SD).

The higher sparseness of the unpaired DES-evoked synergies led us to wonder whether their combinations might provide better explanations for some of the observed behavioral synergies. We subsequently systematically assessed whether synergy vectors resulting from the merging or fractionation of the DES-evoked synergies may be even more similar to the behavioral synergies (Cheung et al., 2012). At the level of the synergy clusters, merging the DES-evoked cluster centroids produced synergy vectors that matched very well with the behavioral cluster centroids (SP of 0.99 ± 0.01, mean ± SD) (Figure 2C) while synergy fractionation did not improve similarity of the matched synergy pairs (see Methods, “Merging and fractionation of DES-evoked muscle synergies”). At the level of individual subjects, if we designate synergy pairs with SP > 0.7 as well-matched pairs (see Methods, “Muscles synergy similarity”), across all subjects 59% of the pre- and post-op behavioral synergies could be well-matched directly to a DES-evoked synergy of the same subject with high similarity (SP of 0.87 ± 0.08, mean ± SD), 10% could be well matched to a synergy produced by merging multiple DES-evoked muscle synergies (SP of 0.85 ± 0.05), and 31% could not be well explained by either direct matching or merging (SP of 0.38 ± 0.22, SP from either direct matching or merging, whichever was higher) (Figure 2D).

### Factors contributing to the differences between DES-evoked and behavioral muscle synergies

The behavioral muscle synergies that could not be matched to DES-evoked synergies or their combinations might indicate that some behavioral synergistic muscle responses are not represented by muscle synergies accessible from the M1, or they may arise from limitations in the experimental design. To determine the degree that experimental study design caused this mismatch, correlations between the behavioral-*versus*-DES-evoked synergy set similarity and subject-demographic variables and other experimental parameters (Table S1) were evaluated. For each subject, synergy set similarity was quantified by first fitting the DES-evoked synergies to the pre-op EMGs of all tasks, and then calculating the R^2^ of this cross-fit (Figure 3A). For demographic parameters, across subjects there was no significant correlation between the cross-fit R^2^ and subject age (Figure S4A) or sex (Figure S4B) even though motor cortical excitability and connectivity are known to be age- and sex-dependent (Ferreri et al., 2017; Perciavalle et al., 2010; Shibuya, et al., 2016). For parameters related to the neurosurgical procedure, the cross-fit R^2^ correlated positively with the number of positive loci identified, but negatively with the mean DES current delivered to the M1 (Figure 3C left, number of positive loci, r = 0.54, p = 0.0566; Figure 3C right, stimulation current, r = -0.61, p = 0.0274).

**Figure 3.**
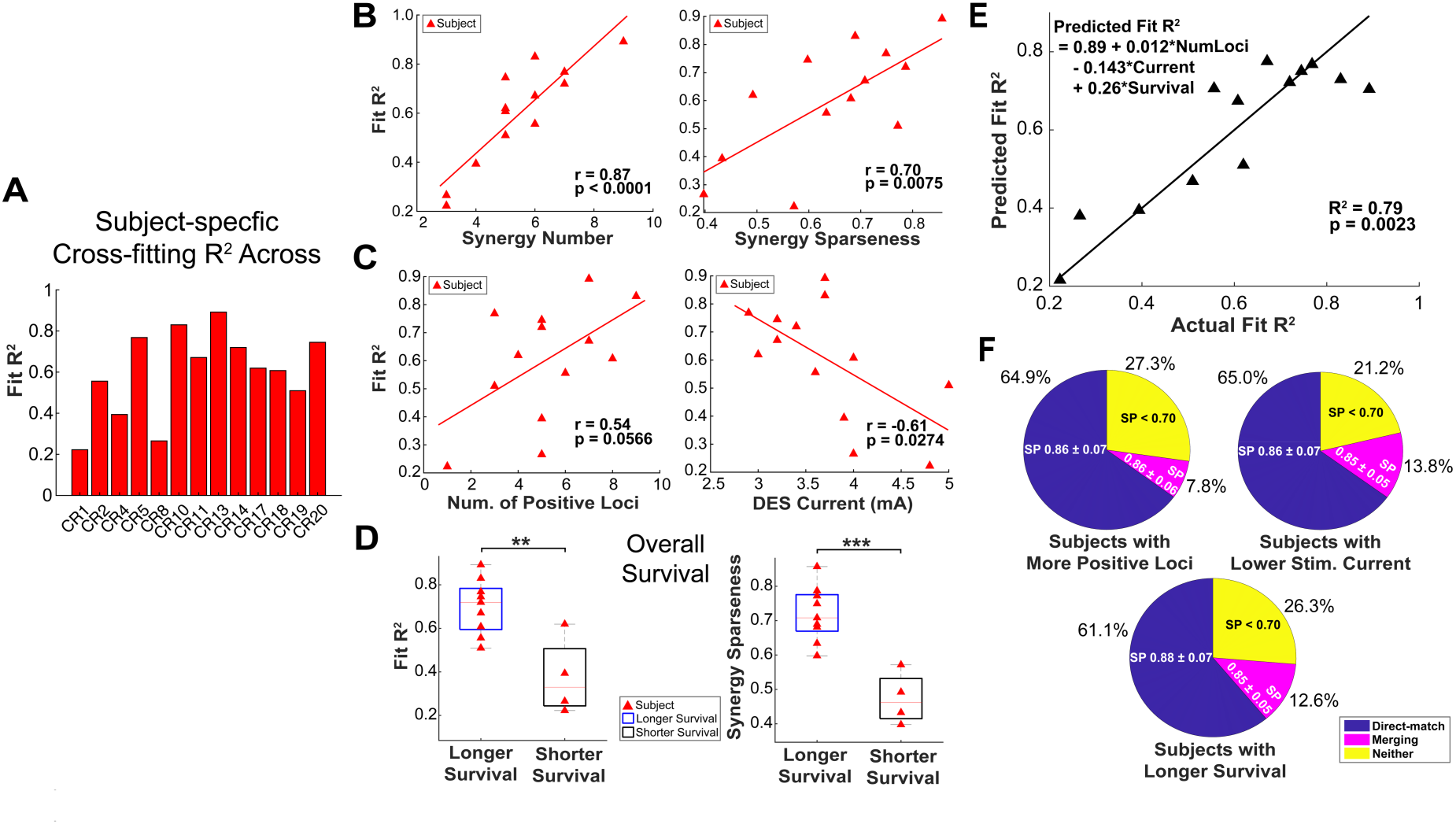
Demographic and experimental parameters influenced the quality of match between the behavioral and DES-evoked muscle synergies. **(A)** The within-subject similarity of DES-evoked and behavioral (pre-op) muscle synergy sets were evaluated by fitting the DES muscle synergies to the behavioral EMGs. Across subjects, this cross-fitting R^2^ showed considerable variability (from ∼0.2 to 0.9). **(B)** Across subjects, both the number of DES muscle synergies extracted (left panel) and average sparseness of the DES synergies (right panel) correlated positively with the DES-to-behavioral cross-fit R^2^. Each red triangle represents data from a subject. The Pearson’s correlation coefficient (r) and its *p* value are shown on each panel. Solid red line denotes a significant relationship (*p* < 0.05). **(C)** Across subjects, the number of positive loci (left panel) and the mean DES current utilized (right panel) correlated significantly with the DES-to-behavioral cross-fit R^2^. Each red triangle represents data from a subject. **(D)** Patient groups with different post-op overall survival showed differences in both the DES-to-behavioral cross-fit R^2^ (left panel) and the average sparseness of the DES-evoked muscle synergies (right panel) (**: *p* < 0.01; ***: *p* < 0.001, independent t-test). Subjects who completed the post-op session after recovery were classified into the longer survival group, and otherwise into the shorter survival group. **(E)** Each subject’s cross-fit R^2^ was predicted with a multiple linear regression of the R^2^ on the number of positive loci (“NumLoci”), average stimulation current (“Current”), and post-op survival time (“Survival”). Each black triangle represents data from a subject. Solid black line describes the linear fit of the regression. **(F)** The percentage of behavioral muscle synergies that could not be accounted for by direct matching to a DES synergy or by merging multiple DES synergies (“Neither” portion in pie charts) was smaller in subgroups of subjects with more positive loci (≥5; top left), lower stimulation current (≤3.7 mA; top right), or longer survival time (bottom).

An additional factor that may influence synergy set similarity is the subjects’ post-op overall survival, which is related to the clinical effectiveness of the neurosurgical procedure (Woo et al., 2023). Subjects were arranged to undergo post-op EMG recording sessions only after they had fully recovered from the surgery. However, some of our subjects (n = 4, 31% of the subjects) did not have sufficient survival time to complete the requisite EMG data collection due to the aggressiveness of their tumor. We therefore classified our subjects into the overall “longer survival” and “shorter survival” groups with the post-op session conducted only in the former. Compared with the shorter survival group, the longer survival group showed a significantly higher cross-fit R^2^ (Figure 3D left, p = 0.003, independent t-test), thus indicating a higher DES-to-behavioral synergy similarity. The World Health Organization (WHO) grades are widely used clinical classifications for brain tumors, which suggest their pathology and underscore prognostic values (Stupp et al., 2014). We thus explored whether the WHO grades of brain tumors, II, III or IV, affected synergy set similarity of our subjects. No significant differences were found in the cross-fit R^2^ among the three WHO grades (Figure S4C top, p = 0.1390, Kruskal-Wallis test). In addition, we investigated whether the type of antiepileptic medications prescribed to the subjects, i.e. valproate, levetiracetam, or phenytoin, influenced synergy similarity as they may affect cortical excitability (Darmani et al., 2019). No significant differences in the cross-fit R^2^ were observed among those treated with any of these medications, and between those with and without such prescriptions (Figure S4D top, p = 0.4003, Kruskal-Wallis test).

To further validate whether the aforementioned factors examined above may influence the DES-to-behavioral synergy set similarity, we used a multiple linear regression model based on the above factors to predict each subject’s cross-fit R². Across subjects, the R^2^ was well predicted by the number of positive loci (“NumLoci”), average stimulation current (“Current”), and the post-op survival time (“Survival”) (reconstruction R^2^ = 0.79, p = 0.0023; Figure 3E). Certain subject subgroups had more positive loci (“NumLoci” ≥ 5, n = 9; determined from mean ± SD of “NumLoci”, 5.23 ± 2.20), required a lower mean DES current (“Current” ≤ 3.7 mA, n = 8; mean ± SD of Current, 3.72 ± 0.64), or had a longer-than-expected survival time (“Survival”, n = 9), and the percentage of their behavioral synergies that could not be matched to DES-evoked synergies or merging of DES-evoked synergies decreased from 30% (Figure 2D) to 27% (“NumLoci”), 26% (“Survival”), and 21% (“Current”), respectively (Figure 3D). In each subgroup, the DES-evoked and behavioral muscle synergy clusters were likewise comparable (Figure S5).

Since the DES-to-behavioral synergy cross-fit R^2^ correlated positively with the number of positive loci (Figure 3C), we theorized that a proportion of subjects had low R^2^ values because the cortical area spanned by their positive loci was not extensive enough to permit the access to all the synergies utilized in the tested motor behaviors. We computationally mimicked this scenario by modelling the unretrieved synergies as an additional synergy extracted from the behavioral EMGs while the fixed DES-evoked synergies were fit to the same EMGs. This extraction-cum-fitting approach was achieved through a manipulation of the NMF algorithm. With this extraction, the cross-fit R^2^ was remarkably improved across subjects (Figure S3B). The additional synergy identified involved mainly upper arm muscles and could be directly matched to at least one of the subject’s behavioral synergies (Figure S3C, S3D). These findings support our interpretation that the unretrieved synergies contributed to the reduced similarity between the behavioral and DES-evoked synergy sets.

Additionally, we observed that across subjects, both the number (Figure 3B, left) and the average sparseness of the DES-evoked muscle synergies (Figure 3B, right) correlated significantly and positively with the DES-to-behavioral cross-fit R^2^ (number of synergies, r = 0.88, p < 0.0001; sparseness, r = 0.70, p = 0.0075). Specifically, the sparseness measure, similar to the cross-fit R^2^, did not vary with the subjects’ age (Figure S4A bottom, p = 0.9699, r = 0.01) or sex (Figure S4B bottom, p = 0.1714, Mann-Whitney U test), nor was it influenced by the WHO grades of brain tumors (Figure S4C bottom, p = 0.2687, Kruskal-Wallis test), nor by treatment with anti-epileptic medications (Figure S4D bottom, p = 0.8346, Kruskal-Wallis test). However, it was higher in the longer overall survival group (Figure 3D right, p = 0.0003, independent t-test). Sparseness of the DES-evoked synergies did not vary with the number of positive loci or stimulation intensity (Figure S4E top, number of positive loci, r = 0.22, p = 0.4719; Figure S4E bottom, stimulation current, r = -0.04, p = 0.8859).

### Reconstructing the cortical map of muscle synergy activity

The results above provide direct physiological evidence that the upper limb muscle synergies for voluntary movement can be independently retrieved through stimulations delivered to the M1. This motivated us to characterize the spatial distribution of the motor cortical loci that activated each muscle synergy, and to examine whether these loci followed a specific or consistent topographical organization. A *sine qua non* for achieving this goal is to obtain the precise anatomical location of each motor cortical locus and reconstruct a standardized brain template that incorporates all stimulation sites across subjects (Viganò et al., 2019; Fornia et al., 2020). Traditionally, the cortical loci stimulated during awake brain surgery are manually marked on structural MRI volumes by the neurosurgeon or referenced with precise MRI coordinates obtained via an intraoperative neuro-navigation system. In our study, the MRI coordinates of the loci were neither acquired during operation nor retained afterwards. To overcome this limitation and achieve greater precision than traditional manual methods, we developed a multi-platform reconstruction pipeline (see Methods, “Reconstructing the map of stimulation loci from MRI”) to systematically map each subject’s stimulated motor cortical loci (Figure 4A). Subsequently, a group brain template was generated by co-registering all stimulation loci of all subjects embedded within the reconstructed cortical surfaces to the left hemisphere (Figure 4B). With this group brain template, we then reconstructed the synergy activation map for each DES-evoked muscle synergy cluster (Figure 2B) by assigning each time- and trial-averaged normalized coefficient of each synergy in the cluster to the location of the trial’s corresponding stimulation locus on the template (Figure 5).

**Figure 4.**
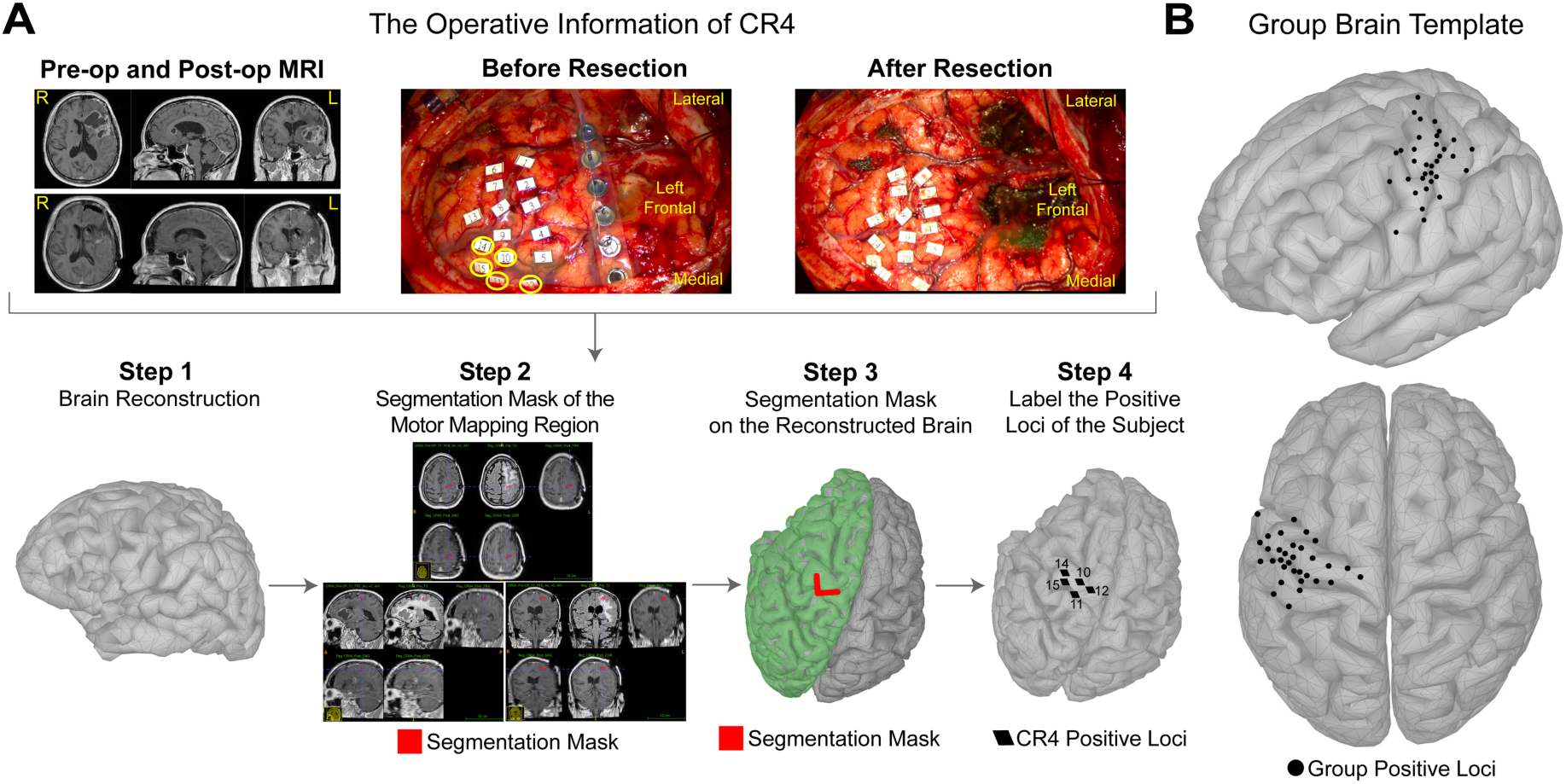
A pipeline for reconstructing the anatomical locations of the stimulated cortical loci of each subject. **(A)** We reconstructed the exact anatomical locations of each subject’s stimulated loci in 4 steps. Step 1: The raw pre-and post-op structural MRI images were converted into 3D brain volumes by MRIcroGL. Then, we reconstructed each subject’s cortical surface from the pre-op T1 volume using FreeSurfer. Step 2: After registering the other pre- and post-op T1/T2 volumes to the pre-op T1 volume, segmentation of the cortical regions mapped by stimulation was conducted in ITK-SNAP based on the registered brain volumes. In the segmentation, we took into account multiple channels of data available, including videos and photos of the field of the surgical microscope recorded during the operation, the planned exposed brain region that was marked on the subject’s scalp under neuro-navigation, the pre-op tumor-affected and post-op resected regions as indicated by MRI etc. Step 3: The segmentation mask from step 2, serving as a user-defined region-of-interest volume, was converted to a cortical surface mask by FreeSurfer (highlighted in red). Step 4: The surface mask from step 3 was mapped to the subject’s reconstructed cortical surface in the Brainstorm toolbox to refine the estimation of the precise locations of the loci. The locations of the positive loci were then manually labeled on the subject’s reconstructed left-side cortical surface, with the location of the surface mask serving as a reference for the labelling. The loci that were originally on the right hemisphere were flipped to the left for this labelling. **(B)** The group brain template co-registered by the cortical surfaces of all subjects to the FSAverage atlas. A total of 68 loci (including 26 right-sided loci projected to the left) were mapped to 35 unique vertices on the left hemisphere of the group template.

**Figure 5.**
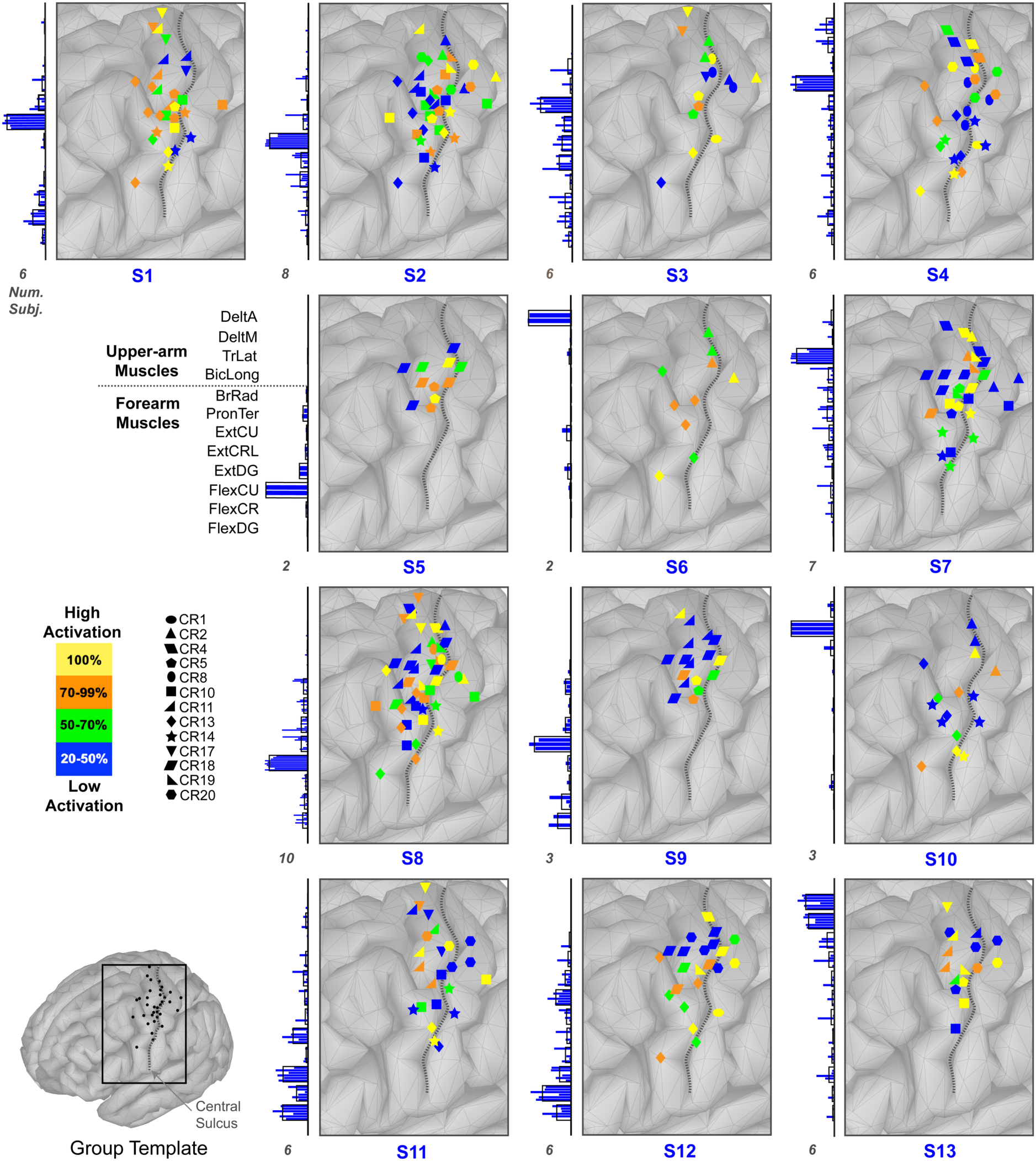
Cortical activity maps of the DES-evoked muscle synergy clusters. For each cluster, the amplitude of each synergy activation coefficient was expressed as a percentage of the maximum coefficient value observed across all positive loci of the subject, with 100% meaning the maximum value. For ease of map visualization, these percentages were categorized into four levels, depicted above with four distinct colors: 100% (yellow), 70-99% (orange), 50-70% (green), and 20-50% (blue). Data from different subjects (CR1 to CR20) are represented by markers with different shapes. In each map, only the cortical tissues in the vicinity of the central sulcus (black dotted line) are shown, with the extent shown demarcated by a rectangle in the brain template shown as an inset above. The map’s corresponding muscle synergy is shown on the left of each map. The number of subjects represented in each cluster is indicated below each muscle synergy plot.

From the reconstructed synergy activity maps (Figure 5), we found that the cortical distribution of the positive loci of the different DES-evoked synergy clusters overlapped extensively. Some of the clusters appeared to share a similar cortical distribution, such as the two extensor-dominated clusters (Figure 5, S2 and S8), the three flexor-dominated clusters (S5, S11, S12), the two clusters with antagonistic forearm muscles (S5 and S9), and the clusters with upper arm muscles (S6, S7, S10). For some of the synergy clusters, the extent of distribution of the loci appeared to be related to the number of muscles activated in the synergy. For example, cluster S1 (from 6 subjects) featured fewer activated muscles and included highly activated loci that spread over a relatively large cortical area; cluster S3 (6 subjects), however, featured a high number of coactivated muscles but exhibited a more localized topographical representation. DES-evoked clusters with a greater number of active muscles, such as S3 and S4, tended to show high-activity loci that distributed along the posterior bank of M1.

### Sparse muscle synergies had more distributed and anterior activity representations in M1

To systematically analyze the cortical maps of synergy activity presented above, we re-represented each map as a 2D matrix, depicted as a heatmap, so that each row of the matrix corresponds to a sequence of antero-posteriorly oriented loci neighboring each other, and each column, a sequence of medio-laterally oriented loci (Figure 6A). For each map, we further calculated the 2D location of the map’s center of mass (CoM) to facilitate comparisons across maps (Figure 6B, red dot). Using the Pearson correlation coefficient to quantify similarity between the cortical maps in their matrix representations, we identified the following groups of DES-evoked synergy clusters, each of which comprises clusters that have similar cortical maps (Table S2): (1) Clusters S6, S7, S10, dominated by upper arm muscles; (2) S3 and S4, dominated by muscles involved in elbow movement; (3) S2 and S8, dominated by forearm extensors; (4) S5, S11, S12, dominated by forearm flexors; (5) S5 and S9, featuring antagonistic muscles; and (6) S7 and S8, as well as S1 and S11, involving functional co-activated muscles for upper limb movement. Thus, muscle synergy clusters with functionally and/or anatomically similar muscle groups also appear to share a similar cortical activity map. Interestingly, the synergies within three of the above cluster groups (S6, S7, and S10; S2 and S8; S11 and S12) may be merged to account for different behavioral muscle synergy clusters (Figure 2C).

**Figure 6.**
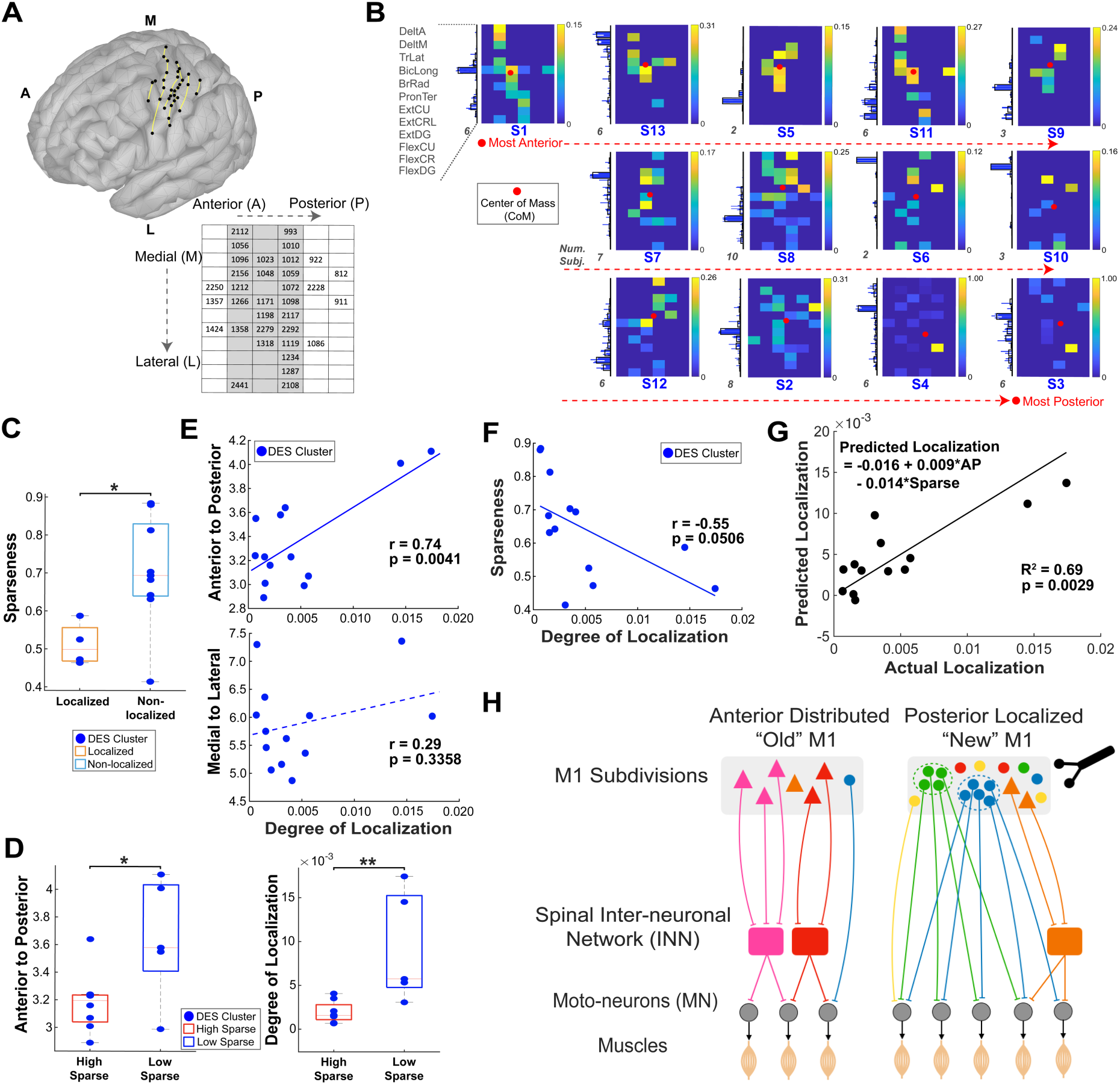
Two subdivisions of M1 with distinct muscle synergy organizational patterns. **(A)** All unique vertices assigned with stimulation loci in the group brain template (N=35) were arranged into a matrix, so that each matrix column corresponds to a mediolateral sequence of neighboring loci, and each row, an anteroposterior sequence. The three matrix columns colored in grey correspond to positions within M1. **(B)** The cortical activity maps of the DES-evoked muscle synergy clusters depicted as heatmaps whose coordinates match the loci positions in the stimulation loci matrix in (A). The 13 maps are ordered here by the anteroposterior position of the map’s center-of-mass (CoM; red dot), with the leftmost map on top (cluster S1) having the most anterior CoM, and the rightmost map at bottom (S3), the most posterior CoM. The anterior maps tend to have a more distributed representation while the posterior maps, a more localized representation. **(C)** DES-evoked synergy clusters with localized representations showed a significantly lower synergy sparseness compared with the non-localized DES clusters. *: *p* < 0.01, independent t-test. **(D)** The high-sparseness DES clusters tended to have more anteriorly distributed cortical maps, as indicated by the CoM anteroposterior coordinate (left panel; *: *p* < 0.01, independent t-test), and lower degrees of activity localization in their representations, as indicated by the map’s activity variance (right panel, **: *p* < 0.001, independent t-test), compared with the low-sparseness DES clusters. **(E)** Across the DES synergy clusters, the map’s degree of activity localization correlated significantly with the anteroposterior position of the map’s CoM (top), but not with mediolateral position of the map’s CoM (bottom). Each blue dot represents data from a DES-evoked muscle synergy cluster. The Pearson’s correlation coefficient (r) and its *p* value are shown on each correlation panel, with solid blue line denoting a significant relationship (*p* < 0.05) and dashed blue line, an insignificant relationship (*p* > 0.05). **(F)** A significant negative correlation between synergy sparseness and the map’s degree of localization was also observed (*p* = 0.0506). **(G)** The degree of activity localization of the clusters’ synergy activity maps could be predicted by a multiple linear regression on the maps’ anteroposterior CoM position (“AP”) and the clusters’ average synergy sparseness (“Sparse”). **(H)** A hypothetical schematic illustrating the two subdivisions of M1 with distinct muscle synergy organizational patterns, as indicated by our results. The anterior subdivision, perhaps corresponding to the “Old M1” of the monkey, contains intermingled and distributed corticospinal populations that coordinate small groups of muscles via the spinal interneurons. The posterior subdivision (“New M1” of the monkey) contains corticomotoneuronal cells for different muscles that are locally bundled together for the coordination of larger muscle groups.

We next explored whether specific features of the cortical maps (such as the maps’ CoM locations) may be related to distinct characteristics of the maps’ corresponding muscle synergies (such as the synergies’ sparseness). We first quantified the pairwise similarity between the cortical maps that shared a common map characteristic, or whose synergies shared a common feature, using three similarity measures: dot product, for evaluating magnitude-sensitive overlap; cosine angle, for evaluating pattern alignment irrespective of magnitude; and Pearson correlation coefficient, for detecting mean-centered linear co-variation (Figure S6 and Table S2). First, similar cortical maps were observed for the subset of synergy clusters that showed high sparseness of the synergy cluster centroid (>0.6; clusters S1, S2, S7, S8, S9, S5, S6, S10; cosine angle of high-sparseness pairs *vs.* all other pairs, p = 0.0098, independent t-test; correlation coefficient, p = 0.0349, Mann-Whitney U test; Fig. S6D), or whose cortical maps were more anteriorly distributed (x-coordinate of CoM < 3.2, clusters S1, S13, S5, S11, S9; dot product of anterior pairs *vs.* all other pairs, p = 0.0249, Mann-Whitney U test; cosine angle, p = 0.00096, independent t-test; correlation coefficient, p = 0.000069, independent t-test; Fig. S6G). When we *k*-means-clustered the cortical maps, one cluster (S3, S4, S11, S13) in particular was composed of maps with higher variance of synergy activations across the stimulation loci (variance of 0.0053 to 0.0174; variance of other maps: 6.55×10^-4^ to 0.0041), which suggested a more localized cortical representation. The muscle synergy centroids associated with these four cortical maps also exhibited significantly lower sparseness as compared with those of the other cortical maps with non-localized representations (p = 0.0285, independent t-test; Figure 6C). Similarly, synergy centroids with higher sparseness also showed cortical maps with less localized cortical distribution (as indicated by smaller variance of synergy activations across the map’s loci) when compared with those with lower sparseness (p = 0.0081, independent t-test; Figure 6D, right). Interestingly, these high-sparseness muscle synergies also had maps with more anterior CoM (p = 0.0278, independent t-test; Figure 6D, left).

These observations prompted us to systematically explore the potential relationships between the degree of activity localization of the cortical map, the CoM location of the map, and the sparseness of the map’s corresponding muscle synergy. Across maps, there was a significant positive correlation between the map’s variance of synergy activations and the x-coordinate of the map’s CoM position (r = 0.74, p = 0.0041, Figure 6E, top). This indicates that the DES-evoked synergy clusters with a more localized cortical activity organization were represented more posteriorly in the M1, and that those with a more uniform distribution were represented more anteriorly. The map’s mediolateral CoM position did not correlate with the map’s degree of localization (r = 0.29, p = 0.3358, Figure 6E, bottom). Also, there was a negative correlation between the sparseness of the synergy centroid and the activation variance of the cortical map (r = -0.55, p = 0.0506, Figure 6F), indicating that the DES-evoked synergy clusters with fewer active muscle components tended to have a more distributed representation across the stimulated loci. An even stronger correlation was obtained when two outliers (from S3 and S4, two low-sparseness DES clusters involved in elbow movement) were excluded from this regression (r = -0.72, p = 0.01).

To validate the above correlations, we used multiple linear regression to predict the degree of synergy activity localization of the cortical map using the map’s anteroposterior CoM position and the sparseness of the map’s muscle synergy. This regression predicted the map’s variance of synergy activations with an R^2^ of 0.69 (p = 0.003; Figure 6G). The above results indicate that sparser muscle synergies had more distributed and more anterior activity representations in the M1, while the non-sparse muscle synergies had more localized and more posterior representations in the M1.

## DISCUSSION

In this study, we successfully retrieved the upper limb behavioral muscle synergies independently through DES delivered to M1 loci during awake craniotomy surgery in patients undergoing glioma resection. We demonstrated the feasibility of accessing muscle synergies through M1 stimulations and mapped out their cortical representations over the M1. Overall, our findings provide direct causal electrophysiological evidence supporting the existence of muscle synergies as neuromotor control units in the human CNS.

### Neural basis of muscle synergies revealed by DES applied to M1

Our main objective here is to validate whether the factorization-decomposed behavioral muscle synergies are genuine substrates of human neuromotor control. Our DES, an invasive yet feasible approach of CNS manipulation in humans, provided valuable access to the neurally based motor modules independent of any experimental motor task design. Most pre-and post-op behavioral synergies (59%) could be described well by DES-evoked synergies at both the subject group (Figure 2B) and individual levels (Figure 2D), and some behavioral synergies (10%) could be reconstructed even better by merging multiple paired and/or unpaired DES-evoked synergies (Figure 2C). Overall, the DES-evoked synergies could reproduce the behavioral synergies decomposed by NMF successfully, supporting the neural origin of the observed behavioral muscle synergies in human motor behaviors (Cheung and Seki, 2021; Huffmaster et al., 2017; Overduin et al., 2012; Rana et al., 2015; Saltiel et al., 2001; Yarossi et al., 2022).

The observed proportion of behavioral muscle synergies explicable by DES is generally consistent with those in analogous studies performed in the monkey. In Overduin et al. (2012), 12 of 18 (67%) upper limb synergies from two monkeys were well matched to synergies derived from intracortical micro-stimulation (ICMS) of M1; in Huffmaster et al. (2017), 3 of 5 (60%) synergies for forelimb reaching significantly correlated with ICMS-evoked synergies. More recently, Yarossi et al. (2022) found that 35 of 40 (88%) hand muscle synergies from 8 human subjects could be matched to synergies elicited from applying TMS to the M1 with dot product >0.7 and an average similarity of 0.77 ± 0.09. Even though our observed percentage of DES-explicable behavioral synergies is lower than that of Yarossi et al., which may be due to our lack of exhaustive M1 mapping, our DES-to-behavioral synergy similarity values were higher, probably because DES recruited more localized cortical networks that were more specific to the generation of particular muscle patterns than TMS (Berardelli et al., 1991).

Before combining the pre-and post-op behavioral synergies for comparisons against the DES-evoked synergies, we evaluated the comparability of the pre-and post-op synergies themselves (Figure 2A). Although each pre-op synergy resembled one of the post-op synergies with high similarity, 21% of the post-op synergies could not be directly matched to a pre-op synergy despite the preservation of the subjects’ motor functions after surgery. In addition, we noticed that DES-evoked synergies provided slightly better explanations to the pre-op than to the post-op synergies (Figure S1). Conceivably, these synergy differences may reflect motor cortical remodeling or neuroplasticity following the operation (Nili et al., 2023; Stålnacke et al., 2024).

Either these synergy changes were not sufficient to lead to any noticeable motor functional impairment, or some of these changes were themselves responses that compensated for synergies’ reorganizations or activations directly resulting from the resection of cortical tissues.

### Unexplained behavioral muscle synergies arise from experimental limitations

In our synergy comparisons, 31% of the behavioral muscle synergies could not be well matched to a DES-evoked synergy or a combination of DES-evoked synergies (Figure 2D). To identify the underlying reasons for this, we first fit each subject’s DES-evoked synergies to the behavioral EMG and obtained the R^2^ of cross-fitting as a more sensitive subject-specific measure to assess the similarity between two synergy sets (Berger et al., 2020; Cheung et al., 2024) (Figure 3A). We then examined whether this cross-fitting R^2^ correlated with parameters related to the subjects’ age, sex, tumor pathological grades, usage of different anti-epileptic drugs, number of identified positive loci, intraoperative DES current, or the DES-evoked synergies themselves. We found that higher fitting R^2^ was associated with more positive loci identified from the subject, lower stimulation current used during mapping, and a longer overall survival time of the subject (Figure 3). The greater number of positive loci identified in certain subjects were likely a result of more comprehensive coverage of M1 during mapping due to the peri-Rolandic location of the tumor in these subjects, which should naturally permit the intraoperative access of more behaviorally relevant muscle synergies. Indeed, when we fit the DES-evoked synergies to the behavioral EMG while allowing additional synergies to be extracted concurrently, these synergies resembled the unmatched behavioral synergies (Figure S3), presumably reflecting the behavioral synergies that were not accessed during surgery. The number of M1 loci that can be stimulated depends on the portion of the M1 exposed after craniotomy, which in turn is dictated by clinical necessity (Roux et al., 2018). It is likely that the behavioral muscle synergies that were unaccounted for come partially from the prohibitive lack of exhaustive M1 cortical exposure owing to clinical needs.

The stimulation current utilized (2 to 6 mA) was determined by the minimal intensity needed to evoke involuntary upper limb movement from the subject. The Penfield stimulation paradigm was used (60-Hz biphasic square pulses over 1-3 sec) and is in keeping with state-of-the-art DES awake motor mapping practice (Bello et al., 2008; Rech et al., 2020; Roux et al., 2020; Ojemann and Silbergeld, 1995; Yingling et al., 1999). DES was delivered with a lower frequency and a longer train than some other DES schemes (Bello et al., 2014; Mottolese et al., 2013; Gogos et al., 2020). Long-train, low-frequency micro-stimulation has been frequently utilized in monkey studies to produce motor cortical outputs that closely resemble the natural firing patterns during voluntary movement (Graziano et al., 2002; Huffmaster et al., 2017; Logothetis et al., 2010; McIntyre and Grill, 2022; Overduin et al., 2012). Also, lower current intensities should more likely recruit localized neuronal networks that activate specific muscle synergies (Saltiel et al., 2001) while higher intensities tend to induce more artificial recruitment patterns through hijacking the natural cortical activities, resulting in severe interruption of the balanced state of the motor cortical network activity (Capaday, 2022; Cheney et al., 2013; Huffmaster et al., 2018). This may explain why a lower current level was associated with higher DES-to-behavioral synergy similarity.

Although the subjects involved in this study did not have any noticeable motor impairment before surgery, it is true that the glioma itself may impact motor cortical functions (Krishna et al., 2023; Mirchandani, et al., 2020; Yu et al., 2020) and the network’s potential in altering its functional connectivity following tumor resection (Nili et al., 2023; Stålnacke et al., 2024). Anti-epileptic drugs prescribed to the patients after surgery may also alter the excitability and microstructure of M1 neurons and the corticospinal tract (Darmani et al., 2019). All of these changes, as well as the tumor type, tumor involved brain structures, and the spread of the tumor could ultimately be reflected in the patients’ overall survival (OS) (Woo et al., 2023) and, possibly, in the muscle patterns used for motor behaviors. We observed that in patients with longer OS, the DES-evoked muscle synergies could explain the behavioral EMG with higher cross-fitting R^2^ (Figure 3D; Figure S5). We speculate that the motor cortical areas of those with longer OS underwent less cortical reorganization in response to the growing glioma, which in turn allowed these subjects to rely more on cortically accessible muscle synergies to generate voluntary motor behaviors. Understanding how exactly the tumor may alter the pathological synergy-encoding network and the muscle synergies and how the OS of patients diagnosed with brain tumors can be predicted could be fruitful directions of future research.

In sum, we believe that the 31% of behavioral muscle synergies that could not be adequately explained by DES-evoked synergies likely arose from experimental limitations stemming from the lack of exhaustive M1 motor mapping, the use of relatively higher stimulation intensities, and the possible glioma-induced motor cortical network disruptions. Even though we cannot entirely exclude the possibility that some of these unexplained behavioral muscle synergies represent muscle synergies of non-neural origin, it is unlikely given that across subjects, at least 3 of the 5 clusters of unexplained behavioral synergies were highly similar to a cluster of explained behavioral muscle synergy (Figure S2).

### Two M1 subdivisions with distinct muscle synergy organizational patterns

Beyond demonstrating the neural origin of the behavioral muscle synergies, our recordings from awake craniotomy surgery have allowed us to reconstruct each synergy’s topographical representation over the M1 as an activation map (Viganò et al., 2019) (Figure 4, Figure 5, Figure 6B). We observed that the maps of the synergies exhibited overlapping and intermingling representations across their cortical access points. Also, the synergy clusters that involved similar functional and/or anatomical muscle groups (e.g. upper arm muscles, extensors, flexors) tended to have similar cortical activation maps. These findings are in general consistent with previous observations in monkeys and humans (Catani, 2017; Devanne et al., 2002; Donoghue et al., 1992; Huffmaster et al., 2018; Overduin et al., 2012; Roux et al., 2020; Griffin et al., 2015; Sanes et al., 1995). In addition, we revealed several significant correlations between characteristics of the DES-evoked synergy clusters and their cortical maps (Figure 6E and 6F). Notably, the synergy clusters with localized cortical maps were primarily activated in posterior M1 regions whereas clusters with more distributed representations were activated in anterior M1 regions (Figure 6E). Furthermore, the synergy clusters that were less sparse in their structures tended to have more localized representations (Figure 6F). Thus, applying DES to the anterior M1 activates sparser muscle synergies that are distributed more uniformly over the cortex; applying DES to the posterior M1 activates non-sparse muscle synergies that are more localized in their cortical representations.

It is tempting to associate the anteroposterior M1 subdivisions as the classically described “Old” and “New” M1 previously reported in the monkey (Sanes and Donoghue, 2000; Rathelot and Strick, 2009). As the M1 division that emerged more recently in evolution, the new M1 is situated more posteriorly and has a higher concentration of corticomotoneuronal (CM) cells that activate individual muscles through direct monosynaptic pathways to the motoneurons (Cheney and Fetz, 1984 & 1985; Fetz, 1976; Kuypers, 1981; Omrani et al., 2017; Rathelot and Strick, 2006; Lemon, 2019). The more anterior old M1, the division with a longer phylogenetic history, has fewer CM cells and is predominantly composed of corticospinal neurons that influence multi-muscle activities through polysynaptic connections with the spinal interneurons (Strick et al., 2021). In humans, evidence has likewise suggested the existence of anatomical and functional M1 subdivisions. Within Broadmann area 4, the anterior 4a and posterior 4p subdivisions show distinct cytoarchitecture, density of pyramidal cells, and fMRI activation patterns (Binkofski et al., 2002; Geyer et al., 1996). Our DES-elicited synergy clusters S3 and S4, featuring co-activated upper and forearm muscles involved in elbow movement, had localized cortical representations located in the posterior “4p” subdivision close to the central sulcus, for example. Subdivisions have also been reported in the “hand-knob” area of the M1 (Sharma et al., 2008) in humans, in that DES applied to the caudal M1 was more likely to elicit positive muscle responses, and multi-muscle activities elicited at the rostral M1 tended to be less correlated (Fornia et al., 2020; Viganò et al., 2019).

Our findings demonstrate that the human M1 is composed of two functionally distinct subdivisions, each exhibiting a unique pattern of synergy organization, and perhaps analogous to the old and new subdivisions in the monkey. One possible synergy organizational scheme consistent with previous data and our results is as follows (Figure 6H). In the anterior subdivision, M1 neurons function to drive downstream spinal interneurons that coordinate the muscles within the sparse muscle synergies, and the M1 neuronal populations that drive different synergies also show distributed, overlapping representations, thus allowing them to spatially intermingle with each other. These muscle synergies, due to their sparsity, may implement fundamental biomechanical functions relevant to many motor behaviors. The intermingling of their cortical access points can therefore allow them to be easily assembled and recruited in different combinations for executing diverse motor tasks. In contrast, for the posterior subdivision, each non-sparse muscle synergy is organized by the muscles’ CM cells that are in close proximity (Rathelot and Strick, 2006), and can therefore be conveniently co-activated as a neuronal cluster. Such an organization accounts for the synergy’s localized representation. Some of the non-sparse synergies organized this way may well be acquired during motor development or motor learning for executing task-specific, specialized biomechanical functions. Indeed, the intermingling of the CM cells of different muscles at the posterior M1 facilitates the assembly of novel muscle coordination patterns as a task-specific muscle synergy (Rathelot and Strick, 2006). We note that due to the limitations of our human DES scheme, our electrodes could not penetrate deeply into the posterior M1 buried within the bank of the central sulcus (Park et al., 2001), which may limit the comprehensiveness of the synergy maps reported here. Conceivably, future mapping experiments may improve the spatial resolution and accuracy of M1 subdivisions with stimulations delivered using finer electrodes or with neuronal recordings.

The proposed roles of M1 in the activation and organization of muscle synergies described above are consistent with the many recent demonstrations that M1 neuronal population dynamics may be best modelled as state-space trajectories that either move within a low-dimensional neural manifold space spanned by a small number of neural modes (Churchland et al., 2012; Gallego et al., 2017; Gallego et al., 2018), and/or evolve as dictated by the structure of a dynamic flow field in the neural state space that provides an overall computation for motor function (Churchland and Shenoy, 2024; Russo et al., 2018; Oby et al., 2025). For instance, the neural modes or the flow field may be structured in a way to activate a set of muscle synergies organized within the cortex or spinal cord. The latent dynamics of the neural population may likewise be related to the dynamics of synergy recruitment, and any robust temporal ordering of states along the trajectory (Oby et al., 2025) may indicate a fixed sequence of muscle synergies recruited for a task. However, the exact relationship between M1 neural trajectories and the dynamics of muscle synergy recruitment, however, remains to be understood.

## CONCLUSION

We argue that this study has advanced our understanding of the origin of muscle synergies in humans by utilizing direct electrophysiological evidence obtained from cortical stimulation in patients undergoing glioma resection. To the best of our knowledge, not only is this study likely the first that demonstrates the neural origin of muscle synergies in humans by exploiting a neurosurgical procedure, but it also provides the most detailed topographical description to date of how human muscle synergies may be represented over the M1 while at the same time revealing the two M1 subdivisions with distinct synergy organizations.

In summary, this work addresses a longstanding controversy in motor neuroscience concerning the origin of behavioral muscle synergies. By comparing the DES-evoked and behavioral muscle synergies, we showed that the NMF-decomposed synergies are not mere algorithmic artifacts or reflections of task constraints, but instead, represent genuine neuromotor substrates of control embedded in the human motor system. Our findings may have implications for the future development of target-driven motor rehabilitative interventions and brain-computer interface applications.

## METHODOLOGY

### Subjects

Patients with gliomas, i.e. primary intraparenchymal brain tumors (N=13, age 33-68, 4 females; Suppl. Table 1) clinically indicated for awake craniotomy for intraoperative brain mapping and tumor resection, were recruited. The subject inclusion criteria were: (1) age 18 to 65 years; (2) confirmed histological diagnosis of World Health Organization grades 1 to 4 gliomas, amenable for resection using this neurosurgical technique; (3) anticipated craniotomy exposing the primary motor cortex (M1); (4) preoperative absence of motor functional impairment. Subject exclusion criteria were: (1) preoperative seizures; (2) preoperative motor functional impairment; (3) any medical contra-indications for awake craniotomy; (4) skin allergies to alcohol, metal surfaces, medical tapes or bandages; (5) substantial cognitive impairment, or difficulties in understanding or following verbal instructions in English or Cantonese. The study’s protocol was approved by the Research Ethics Committee/Institutional Review Board (Kowloon Central / Kowloon East Clusters) of the Hong Kong Hospital Authority (protocol no. KC/KE-21-0012/ER-2).

### EMG recordings during voluntary motor tasks

Each subject underwent pre-operative (3-5 days before surgery) data recording, and a post-operative recording session (3-6 months after surgery). Multi-muscle electromyographic (EMG) data were collected as the subject performed upper limb voluntary tasks related to basic activities of daily living (Figure 1A). While pre-operative data were collected from all subjects, post-operative data were collected from the subjects (N=9) who were in good physical condition and had preserved motor function after a period of motor recovery. In both the pre- and post-operative sessions, surface EMGs (2000 Hz) were recorded from 12-15 upper limb muscles contralateral to the side of the tumor. Wireless EMG sensors (Trigno, Delsys Inc., Natick, MA) were attached to skin surface. When determining the positions of electrode attachment, we followed the guidelines for Surface Electromyography for the Non-Invasive Assessment of Muscles — European Community project (SENIAM) (www.seniam.org). The muscles recorded included: infraspinatus (Infrasp); pectoralis major, clavicular head (PectClav); trapezius, superior fibers (“major” head) (TrapMaj); deltoid, anterior (DeltA) and medial parts (DeltM); triceps, lateral head (TrLat); biceps, long head (BicLong); brachioradialis (BrRad); pronator teres (PronTer); extensor carpi ulnaris (ExtCU); extensor digitorum (ExtDG); extensor carpi radialis longus (ExtCRLongus); flexor carpi ulnaris (FlexCU); flexor carpi radialis (FlexCR); and flexor digitorum (FlexDG). All executed motor tasks were also documented with video recordings.

During task execution, subjects sat with their backs straight on a chair without armrest support. In each session, the subject was instructed to perform 10-11 upper limb behavioral tasks (5 trials for each), including reaching without elbow extension, reaching with elbow extension, shoulder abduction, forearm and wrist pronation, shoulder circumduction, elbow flexion, fist opening and closing, concurrent arm and hand opening, grasping, gripping, and screw-driving. These tasks were previously used to study muscle coordination during upper limb voluntary movement (Cheung et al., 2009, 2012).

### Awake craniotomy brain surgery

Before the operation, structural magnetic resonance imaging (MRI) (T1, T2, T2-FLAIR sequences) of the subject’s brain were acquired. The pre-operative MRI scans were reviewed to demarcate the cortical areas to be exposed by the craniotomy. On the day of surgery, wireless EMG sensors were attached to the subject’s skin to collect intraoperative EMG data. To prevent any discomfort that may arise when the subject is positioned intraoperatively by the neurosurgeon for surgical access, sensors for the muscles Infrasp, PectClav, and TrapMaj were typically not recorded.

The awake craniotomy was performed according to the standard asleep-awake-asleep approach whereby monitored anesthetic care was delivered by total intravenous anesthesia using propofol and remifentanil (Min, 2025). Bispectral index monitoring was adopted to assess the subject’s depth of sedation. The subject was not administered any muscle relaxing agent throughout the procedure. Upon initiating intraoperative brain mapping, all sedative medication was ceased and the patient was allowed to return to full consciousness. Direct electrical stimulation of the cortex was performed using the Penfield technique (Hervey-Jumper et al., 2015; Penfield and Boldrey, 1937). A bipolar electrode delivered a constant current beginning with 2 mA and increased to a maximum of 6 mA until somatosensory or motor function was detected at the cortical region of interest. The current was generated with 1.25-msec biphasic square waves in 1-3 second trains at 60 Hz (Ojemann stimulator, Radionics, Burlington, MA, USA). The stimulation ball-tip electrodes were each 1 mm in diameter and separated 5 mm apart.

When the subject returned to full consciousness, continuous EMG monitoring, video recording of the subject’s arm movement, and video capturing of the operating field were initiated. Mapped cortical regions, including M1, were registered using a neuro-navigation system (Brainlab^®^, Munich, Germany) and also indicated using cortical markers (Figure 1A). The subject then began performing a series of preoperatively defined repetitive voluntary upper limb tasks, grasping, gripping, and elbow flexion at a self-selected pace. The threshold amplitude of the stimulation current needed for each subject’s motor mapping was determined by gradually increasing or decreasing the current from 2 mA until involuntary muscle activities were evoked in at least one locus while ensuring that no seizures or abnormal electrocorticographic (ECoG) after-discharges were induced (Bello et al, 2008). If a motor task was interrupted or stopped by stimulation-evoked involuntary muscle contractions when stimulation was being delivered to a locus, that locus would be marked as a positive locus (i.e., one that yielded a positive motor response) (Penfield and Boldrey, 1937). Typically, to define the threshold stimulation intensity, the current amplitude was adjusted over 3 rounds of stimulation involving 10-20 loci.

A further 1 to 3 additional rounds of stimulation were conducted over 10-20 minutes on the identified positive loci to permit the collection of additional stimulation-derived EMGs. During this phase of recording, the subject was instructed either to relax without performing any tasks, or to perform repeated grasping, gripping, or elbow flexion motor tasks as before when stimulation was delivered at threshold intensity. For the majority of the subjects (N=10), currents at supra- and/or sub-threshold intensities were also tested over 1 to 3 rounds (Feinsinger et al., 2022).

A post-operative MRI of the same sequences was acquired the next day. The pre- and post-operative images were compared to localize as well as to confirm the extent of tumor and brain tissue resection.

### EMG preprocessing

For the data collected from the pre- and post-op behavioral sessions, each channel and each episode of raw EMG data were carefully inspected visually for evaluation of signal quality. The channels or episodes with poor signal quality or generated from incorrect motor tasks were omitted from further analysis. Each EMG channel was preprocessed with high-pass filtering (window-based finite impulse response [FIR] filter, cut-off frequency of 50 Hz), rectification, low-pass filtering (window-based FIR, cut-off frequency of 20 Hz), baseline noise removal, 20-ms integration, and spike noise removal, following the steps described in previous studies (Cheung et al., 2020; Cheung et al., 2024). Subsequently, each EMG channel of each session (pre- or post-op) was normalized to the maximum EMG amplitude observed in the channel’s corresponding muscle in that session.

For the EMG data collected in the intraoperative session, based on careful visual inspection and temporal alignment of the EMG traces, the video of the subject’s arm movement, and the video of the surgical field, each continuously recorded raw EMG trace was parsed into segments, with each segment containing the DES-evoked muscle activities from a stimulation trial of a positive locus. The number label of the segment’s corresponding stimulation locus and the stimulation current employed in that trial were also noted (Figure 1B). After the isolation of all DES-evoked motor responses, each EMG channel was preprocessed with the same steps as described above except that the last spike noise removal step was not executed.

### Extraction of muscle synergies

The non-negative matrix factorization (NMF) algorithm (Lee and Seung, 1999) was used to model the EMG activities and decompose all preprocessed EMGs into a linear combination of time-invariant muscle synergy vectors scaled by time-varying activation coefficients. The NMF model can be expressed by the following equation:

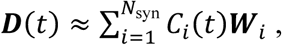

where **D**(*t*) is a matrix of the collected EMG activities over time *t*, C*_i_*(*t*) is the nonnegative time-varying activation coefficients for the *i*^th^ synergy, **W***_i_* is the *i*^th^ nonnegative muscle synergy vector, and N_syn_ is the number of extracted muscle synergies.

For the pre- and post-op behavioral EMG datasets, to determine the appropriate number of the extracted synergies, for each tested number of synergies (from 1 to the number of muscles in the data set), synergy extraction was repeated 20 times, each time with different initial estimates of C*_i_*(*t*) and **W***_i_*. Then, the minimum number of synergies that could describe the data with an EMG-reconstruction R^2^ of ≥80% in at least one of its extraction repetitions was regarded as the “best” number of muscle synergies for the dataset. In each extraction, convergence was defined as a change of EMG-reconstruction R^2^ < 0.001% in two consecutive iterations (Cheung et al., 2012).

For the intraoperative DES-evoked EMGs, considering that the number of data points available for synergy extraction in each dataset was always smaller than those in the behavioral datasets, the number of synergy extraction repetitions was increased to 50 for each tested synergy number, and the EMG-reconstruction R^2^ threshold for selecting the number of synergies was likewise increased to 90% (Fricke et al., 2020; Huffmaster et al., 2017; Yarossi et al., 2022). To further assess the robustness and generalizability of the muscle synergies extracted from the DES-evoked EMG, in each dataset we implemented an additional cross-validation step for 50 times, each time by extracting synergies from 50% of the EMG data points, randomly selected, and then fitting the extracted synergies to the remaining data for testing.

After extracting the DES-evoked muscle synergies from the EMGs, for each synergy we averaged the activation coefficient across the duration of each trial, and then further computed the average activation coefficient value across all trials of each stimulation locus. These average coefficient values for the different loci were then normalized to the maximum coefficient value for that synergy observed after averaging. With these normalized average coefficient values, we then generated the synergy’s cortical activation map by displaying these values in a 2D heatmap whose coordinates correspond to the anatomical locations of the stimulation loci on the cortical surface (see below) (Huffmaster et al., 2017).

### Sparseness of muscle synergies

When inspecting the muscle synergy vectors extracted from the behavioral or DES-evoked EMGs, we found it useful to quantify the number of active muscle components in each muscle synergy vector as the vector’s sparseness. We adopted the definition proposed by Hoyer (2004) to quantify the sparseness of each muscle synergy:

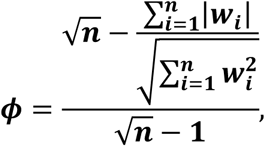

where φ is the vector’s sparseness index (with φ = 1 meaning that the vector has a single non-zero active component, and φ = 0 meaning that the all components in the vector are equally activated), n is the number of muscle components in the muscle synergy, and W*_i_* denotes the amplitude of the *i*^th^ muscle component of the nonnegative muscle synergy vector **W**.

### Clustering muscle synergies

The behavioral and DES-evoked muscle synergies of all subjects were respectively categorized into synergy clusters by *k*-means clustering. Each run of clustering was repeated 1000 times, each time with different estimates of the initial centroids of the clusters. Clustering was initially performed with 2 to 20 clusters. The optimal number of clusters was determined either by identifying the cluster number that yielded a robust local maximum of silhouette value for the synergy clusters of the pre- and post-op behavioral sessions, or by the gap statistics method (Cheung et al., 2020; Tibshirani et al., 2001) for the intraoperative stimulation sessions. A different method was used for the DES-evoked synergies because this synergy set contained more extreme data points, and the gap statistics method is known to yield more robust estimates for such a dataset.

### Muscle synergy similarity

To assess the similarity between two muscle synergy vectors (e.g., from the pre- and intraoperative sessions) or the centroids of two synergy clusters, the scalar product was calculated after the vectors were L2-normalized (Cheung et al., 2009). In our analysis, we found it useful to classify vector pairs with scalar product >0.7 as synergy pairs with higher similarity (d’Avella et al., 2003; d’Avella et al., 2006; Cheung et al., 2009). This threshold was determined based on the average scalar product value of all DES-to-behavioral best-matching synergy pairs of all subjects (0.6959 ± 0.28, mean ± SD).

To further compare the DES-evoked intraoperative muscle synergies with the pre- and post-op behavioral synergies, we also adopted a more holistic approach of comparison by assessing how well the synergy set from one condition may describe the EMG of another condition. For each subject, the DES-evoked synergies were fit, as a “baseline” synergy set, to different sets of behavioral muscle synergies using the NMF algorithm. Fitting was done by holding the synergy vectors constant while updating the time coefficients across iterations as the algorithm was run (Berger et al., 2020; Cheung, 2020). The quality of fit was then quantified by the EMG reconstruction R^2^ value. The larger the R^2^, the better the baseline synergies can generalize to describing the new dataset. To explore whether extra synergies in addition to the baseline set could substantially increase the R^2^ of the cross-fit, we manipulated the NMF algorithm by allowing extra new muscle synergies (N=1) to be extracted concurrently from the behavioral EMGs while the fixed DES-evoked synergies were being fit to the data (Figure S3). If the overall cross-fit R^2^ increases with the extraction of these additional synergies and that the extra synergies extracted resemble some of the original behavioral synergies, we may interpret the extra muscle synergies to reflect either synergies that are missed and therefore not retrieved by the stimulation, or muscle coordination patterns that contribute to the motor behavior but are not accessible from the motor cortex.

Task-specific behavioral muscle synergies of each subject were also extracted from the pre-op EMG of each of the 11 upper limb tasks. Then, the DES-evoked muscle synergies were matched to these task-specific synergies, and the scalar product values of the matched synergy pairs were obtained for each task and each subject (Figure S3A). Furthermore, we fit the DES-evoked synergies to the EMG of each task and calculated the R^2^ of the fit to assess whether the DES-evoked patterns can best explain the EMGs of a subset of the tasks (Figure S3B).

### Merging and fractionation of DES-evoked muscle synergies

Previous studies have shown that new muscle synergies may be accounted for either by merging multiple pre-existing synergies together, or by fractionating a pre-existing synergy into multiple smaller modules (Cheung et al., 2012; Cheung et al. 2024). Here, we explored whether some of the behavioral muscle synergies may be explained by merging or fractionating the DES-evoked synergies. Muscle synergy merging may be formalized by the following model (Cheung et al., 2009, 2020):

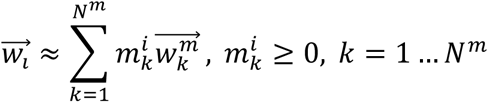

where **w**_i_ is the *i*^th^ behavioral muscle synergy vector, **w**^m^ is the *k*th to-be-merged DES-evoked muscle synergy vector, m^i^ is a nonnegative coefficient, and N^m^ is the number of synergies to be merged. Each behavioral synergy was modeled as a linear combination of a set of DES-evoked synergies by projecting each behavioral synergy vector onto the EMG subspace spanned by the DES-evoked synergies (as basis vectors). The coefficients of this linear combination were obtained by standard nonnegative least squares optimization. Following Cheung et al. (2012), if m^i^ ≥ 0.2, the coefficient’s corresponding **w**^m^ was regarded as a contributor to the merging. Fractionation of a DES-evoked muscle synergy was similarly defined as the converse of the synergy merging model above. The DES-evoked synergy was modeled as a linear combination of a set of behavioral muscle synergies. In this vector projection, we imposed the additional constraint that each behavioral synergy could only be the fractionation of a single DES-evoked synergy (Cheung et al., 2012). The scalar product values between the original behavioral synergy and the behavioral synergy reconstructed from synergy merging or fractionation were then computed. The higher of these two values would determine whether merging or fractionation would be the better model for explaining that behavioral synergy.

### Reconstructing the map of stimulation loci from MRI

To further investigate the spatial distribution of the motor cortical loci whose stimulations activate muscle synergies, it is essential to obtain the precise anatomical locations of the identified positive loci, and then build a brain map that incorporates these loci for each subject. In our study, the 3D coordinates of the stimulated loci were not acquired in real-time during operation. Therefore, we developed a multi-platform pipeline to reconstruct the exact anatomical locations of the stimulated cortical loci of each subject (Figure 4). The accuracy of the pipeline’s output and average distance between loci were marked by two raters under the supervision of the surgeon who performed the craniotomy and executed the stimulation.

In this pipeline, the raw pre- and post-op structural MRI images (DICOM format) of all subjects were first converted into 3D brain volumes (NIfTI format) by a built-in conversion function of MRIcroGL (version 1.2; www.nitrc.org). For each subject, the first step was to reconstruct the subject’s cortical surface from the pre-op high-resolution 3D T1 volume using FreeSurfer (recon-all-clinical function; Billot et al., 2023a, 2023b; Gopinath et al., 2023; Iglesias et al., 2023). After registering the other pre- and post-op T1/T2 volumes to the pre-op T1 used in the previous step (Reuter et al., 2010), segmentation of the cortical regions mapped by stimulation was conducted in ITK-SNAP (version 4.0.1; www.itksnap.org) based on the registered brain volumes (Yushkevich et al., 2006). In the segmentation, we took into account multiple channels of data collected before, during, and after the operation, including videos and photos of the field of the surgical microscope recorded during the operation, the planned exposed brain region (including M1 and those afflicted by the tumor) that was marked on the subject’s scalp under neuro-navigation after the subject’s surgical position was fixed, the motor mapping plan, and the pre-op tumor-affected and post-op resected regions as indicated by MRI. This segmentation mask, serving as a user-defined region-of-interest (ROI) volume, was converted to a cortical surface mask by FreeSurfer.

Subsequently, this surface mask was mapped to the subject’s reconstructed cortical surface in the Brainstorm toolbox (Tadel et al., 2011) to refine the estimation of the precise locations of the stimulated loci. The individual brain template of each subject was reconstructed by 5003 vertices in Brainstorm to ensure that the area spanned by the stimulated loci on the reconstructed brain matches the actual area spanned by the stimulated loci on the subject’s cortex. For each subject, the locations of the positive stimulation loci were then manually labeled on the subject’s reconstructed left-side cortical surface, with the loci that were originally on the right hemisphere flipped to the left for this labelling. We compiled all stimulation sites onto the same hemisphere so that the synergy activation maps of the entire cohort could be constructed from the maximal amount of data available (see below). Finally, a group template with the locations of all positive stimulation sites was generated by co-registering the reconstructed cortex, now labelled with loci, of all subjects in the MNI space using the Brainstorm toolbox (Ashburner and Friston, 2005). A total of 68 loci (including 26 loci projected from the right hemisphere) were mapped on this group template of the left hemisphere.

### Cortical maps of muscle synergy activities

The cortical map of the stimulation loci obtained from the abovementioned steps allowed us to construct another map that depicts how the activation of each muscle synergy varied as different cortical loci were stimulated during the intraoperative session. To generate the synergy activity map of each intraoperative synergy, the synergy’s coefficient for each positive stimulation locus, averaged across the stimulation trial’s duration and then across multiple trials at that locus (see above), was first expressed as a percentage of the maximum coefficient value of that synergy observed across all positive loci of the subject, with 100% meaning the maximum value. These percentage values were then depicted as a heatmap at the locations of their corresponding cortical loci in the map of stimulation loci. To ease visualization, we found it useful to categorize these coefficient percentages into 4 bands: 20-50% (shown in the map in blue), 50-70% (green), 70-99% (orange), and 100% (yellow). The individual activity maps of all muscle synergies within an intraoperative synergy cluster were then combined together to produce an overall map for each cluster with all coefficient percentages depicted in the co-registered 3D brain template for the entire cohort (Figure 5).

To ease subsequent analysis and visualization of the synergy activity maps, we derived an alternative representation of every cortical map by remapping the synergy coefficient data from the 3D brain template to a 2D linear surface. The entire set of left hemispheric cortical locations of the 3D template (Figure 4B), specified by the 3D coordinates of 5003 vertices (same number of vertices in the reconstructed cortex of each individual), was registered to the FSAverage atlas using an inflated spherical representation based on the cortical folding patterns (Fischl et al., 1999; Reuter et al., 2012), and then flattened into a 2D surface in the Brainstorm toolbox. This process yielded the 2D positions of all stimulation loci whose locations spread over the area defined by vertices no. 812 to 2441. In this remapping, we found that all 68 stimulation loci could be mapped to 35 vertices, but the different loci of every subject could always be uniquely mapped to distinct vertices. Thus, an individual vertex on this group brain template may correspond to multiple neighboring stimulation loci with each of which coming from a different subject. For every synergy cluster, the synergy coefficient percentages assigned to these vertices were then conveniently represented as entries within a 12✕6 matrix (containing 72-35 = 37 blank entries), with each matrix column corresponding to a line of medial-to-lateral loci neighboring each other, and each row, a line of anterior-to-posterior loci (Figure 6A, bottom). Coefficient percentage values from different subjects assigned to the same vertex were averaged. Matrix values of each synergy cluster were then visualized as a 2D rectangular heatmap (Figure 6B).

For each synergy activity map, the coordinates of the center of mass (CoM) of the map’s activity distribution were also calculated so that maps with different CoM positions could be compared. Similarity between any two synergy activity maps was evaluated by correlation coefficient as well as the dot product and cosine angle measures after the matrices were rearranged as vectors (Figure S6 and Table S2).

### Statistics

Descriptive statistical analysis was performed to evaluate if significant differences in mean exist across groups or if there are significant correlations between characteristics. The distribution of the samples was first evaluated by the Lilliefors test to test normality. For datasets with normal distributions, the independent t-test was employed for the comparison of two independent groups, and one-way Analysis of Variance (ANOVA) was employed for the comparison of multiple independent groups. For datasets with non-normal distributions, the Mann–Whitney U test was used for two independent groups, and the Kruskal–Wallis test was used for the comparison of multiple independent groups. Pearson’s correlation coefficient was used to assess possible relationships between different characteristics of muscle synergies. All statistical null hypotheses were rejected at a significance level of α = 0.05. Except for neuroimaging analysis, all data analysis was performed in the MATLAB environment (2022b, MathWorks, Natick, MA).

## INFORMATION

Supplemental information can be found online.

## Data Availability

All data produced in the present study are available upon reasonable request to the authors.

## ACKNOWLEDGMENTS

We would like to express our gratitude to the patients and their families for their participation in this study. We sincerely thank all the neurosurgeons, nurses and clinical psychologists at Kwong Wah Hospital for their invaluable assistance in our data collection throughout the study. Special thanks to Jack Man Wui Li for his contributions to the EMG preprocessing, Richard Liu for technical support in the neuroimaging reconstruction, and Fung Ting Kwok for his insightful comments on the manuscript. The work was approved by Hong Kong Hospital Authority Research Ethics Committee (KC/KE-21-0012/ER-2), and supported by Hong Kong Research Grants Council (14114721, 14119022, N_CUHK456/21, R4022-18F), CUHK RSFS (3133184), CUHK Faculty of Medicine Direct Grants (4054652, 4054710).

## AUTHOR CONTRIBUTIONS

V.C.K.C., P.Y.M.W, R.H.M.C, and J.J.X conceived the study; P.Y.M.W performed intraoperative brain mapping and direct electrical stimulation; A.H.S.K. provided intraoperative anesthetic care for patients; J.J.X, S.H. and K.L collected the data; J.J.X and S.H analyzed the data; J.J.X created the analysis pipeline, interpreted the data, and drafted the manuscript; V.C.K.C., P.Y.M.W, R.H.M.C and J.J.X revised the manuscript.

