## Supplementary materials for "Neuromotor Modules Revealed by Intraoperative Direct Electrical Stimulation of the Human Primary Motor Cortex"

**Supplementary Table 1.** Demographic and clinical information of all subjects.

| | **Age (years)** | **Sex** | **Tumor mass location** | **Tumor pathology with WHO grade** | **Number of positive loci** | **Mean DES current (mA)** | **Post-op session (available = 1)** | **Usage of anti-epileptic drugs** |
|---|---|---|---|---|---|---|---|---|
| **CR1** | 60-65 | M | Left temporal lobe | Anaplastic ganglioglioma (III) | 1 | 4.8 | 0 | Valproate |
| **CR2** | 65-70 | F | Left frontoparietal lobe | Glioblastoma, IDH-wide-type (IV) | 6 | 3.6 | 1 | Valproate |
| **CR4** | 55-60 | F | Left frontotemporal lobe | Anaplastic oligodendroglioma (III) | 5 | 3.9 | 0 | Valproate |
| **CR5** | 35-40 | M | Left frontal lobe | Oligodendroglioma, IDH-mutant (II) | 3 | 2.9 | 1 | Levetiracetam |
| **CR8** | 55-60 | M | Right temporal lobe | Glioblastoma, IDH-wide-type (IV) | 5 | 4 | 0 | Levetiracetam |
| **CR10** | 55-60 | F | Left frontal lobe | Glioblastoma (IV) | 9 | 3.7 | 1 | Levetiracetam |
| **CR11** | 45-50 | M | Right temporal lobe | Glioblastoma, IDH-wide-type (IV) | 7 | 3.2 | 1 | Valproate |
| **CR13** | 60-65 | M | Left temporoparietal lobe | Glioblastoma, IDH-wild-type (IV) | 7 | 3.7 | 1 | Valproate |
| **CR14** | 60-65 | M | Left insular cortex | Oligodendroglioma, IDH-mutant (II) | 5 | 3.4 | 1 | Levetiracetam |
| **CR17** | 40-45 | F | Right frontal lobe | Oligodendroglioma, IDH-mutant (II) | 4 | 3 | 0 | Levetiracetam |
| **CR18** | 60-65 | M | Left temporal lobe and insular cortex | Glioblastoma, IDH-wide-type (IV) | 8 | 4 | 1 | Valproate |
| **CR19** | 45-50 | M | Right frontal lobe and insular cortex | Astrocytoma, IDH-mutant (II) | 3 | 5 | 1 | Valproate |
| **CR20** | 50-55 | M | Right frontal lobe | Astrocytoma, IDH-mutant (II) | 5 | 3.2 | 1 | Phenytoin |

Abbreviations: DES, direct electrical stimulation; WHO, World Health Organization; IDH, isocitrate dehydrogenase.

**Supplementary Table 2.** Summary of comparison results for DES-cluster synergy activity maps.

| Features / DES Cluster Subsets | (a) Similarity between cortical activity maps of DES clusters within subset | | | (b) Correlation coefficient of paired DES cortical activity maps within DES subset | (c) Degree of localization of DES-cluster cortical activity maps | (d) CoM of DES-cluster cortical activity maps | | (e) Sparseness of DES clusters |
|---|---|---|---|---|---|---|---|---|
| | Dot product | Cosine angle | Corr. coef. | | | A-P | M-L | |
| **1 Direct-match** | ns | ns | ns | **** 3/10 pairs<br>*** 2/10 pairs<br>** 1/10 pair | ns | ns | ns | ns |
| **2 Merged** | ns | ns | ns | *** S6, S7, S10<br>* S2, S8<br>** S11, S12 | ns | ns | ns | ns |
| **3 High Sparse** | ns | ** | * | **** 8/28 pairs<br>*** 3/28 pairs<br>** 5/28 pairs<br>* 5/28 pairs | * | * | ns | n/a |
| **4(i) Upperarm** | * | ns | ns | ** S6, S7, S10<br>** S3, S4 | ns | ns | ns | ns |
| **(ii) Forearm extensors** | ns | ns | ns | * S2, S8<br>**** S8, S9 | ns | ns | ns | ns |
| **(iii) Forearm flexors** | ns | ns | ns | **** S5, S11<br>*** S5, S12 | ns | ns | ns | ns |
| **5 Localized** | ns | ns | ns | **** S3, S4<br>**** S11, S13 | n/a | ns | ns | ** |
| **6 Anterior** | * | *** | **** | ** S1, S11, S13 | ns | n/a | | ns |

The table summarizes all comparison results for different subsets of DES-evoked muscle synergy clusters. In column (a), similarity between the cortical synergy activity maps of any two clusters within the subset was evaluated by three different measures (dot product, cosine angle, and correlation coefficient). The mean of this similarity was then compared against the pairwise similarity between the maps of all other pairs of clusters (i.e., including pairs of any two clusters outside the subset, and pairs with a within-subset plus an outside-subset cluster). This result is also presented in Supplementary Figure 6. In column (b), we tabulate the number of pairs of synergy clusters within each subset showing a significant correlation coefficient between their cortical synergy activity maps. In columns (c) to (e), the degree of localization of the synergy cortical map (column (c)), the anteroposterior (A-P) or mediolateral (M-L) coordinates of the center-of-mass of the cortical activity map (column (d)), and the sparseness of the muscle synergy cluster centroid (column (e)), for the clusters within the subset were compared against those for the clusters not in the subset. In these comparisons, 6 subsets of muscle synergy clusters were considered: 1, DES-evoked muscle synergy clusters that could be well-matched directly to a behavioral synergy cluster ("Direct-match", clusters S4, S6, S12, S13, S8; see Fig. 2B); 2, Synergy clusters that could be merged with another DES-evoked cluster for explaining a behavioral synergy cluster ("Merged", clusters S3 & S11, S6 & S7 & S10, and S2 & S8; see Fig. 2C); 3, Synergy clusters with high synergy sparseness ("High Sparse", clusters S1, S2, S7, S8, S9, S5, S6, S10; sparseness > 0.6); 4, Synergy clusters that involved upper arm muscles ("Upperarm", clusters S4, S5, S7, S10, S13; see

Fig. 2B); 5, Synergy clusters with localized cortical distributions (“Localized”, clusters S3, S4, S11, S13; cortical map variance of 0.0053 to 0.0174); 6, Synergy clusters with an anterior activity distribution, as indicated by an anterior CoM position (“Anterior”, clusters S1, S13, S5, S11, S9; x-coordinate of CoM < 3.2). *, $p < 0.05$; **, $p < 0.01$, ***, $p < 0.001$, ****, $p < 0.0001$. Abbreviations: CoM, center of mass; n/a, not available; ns, not significant.

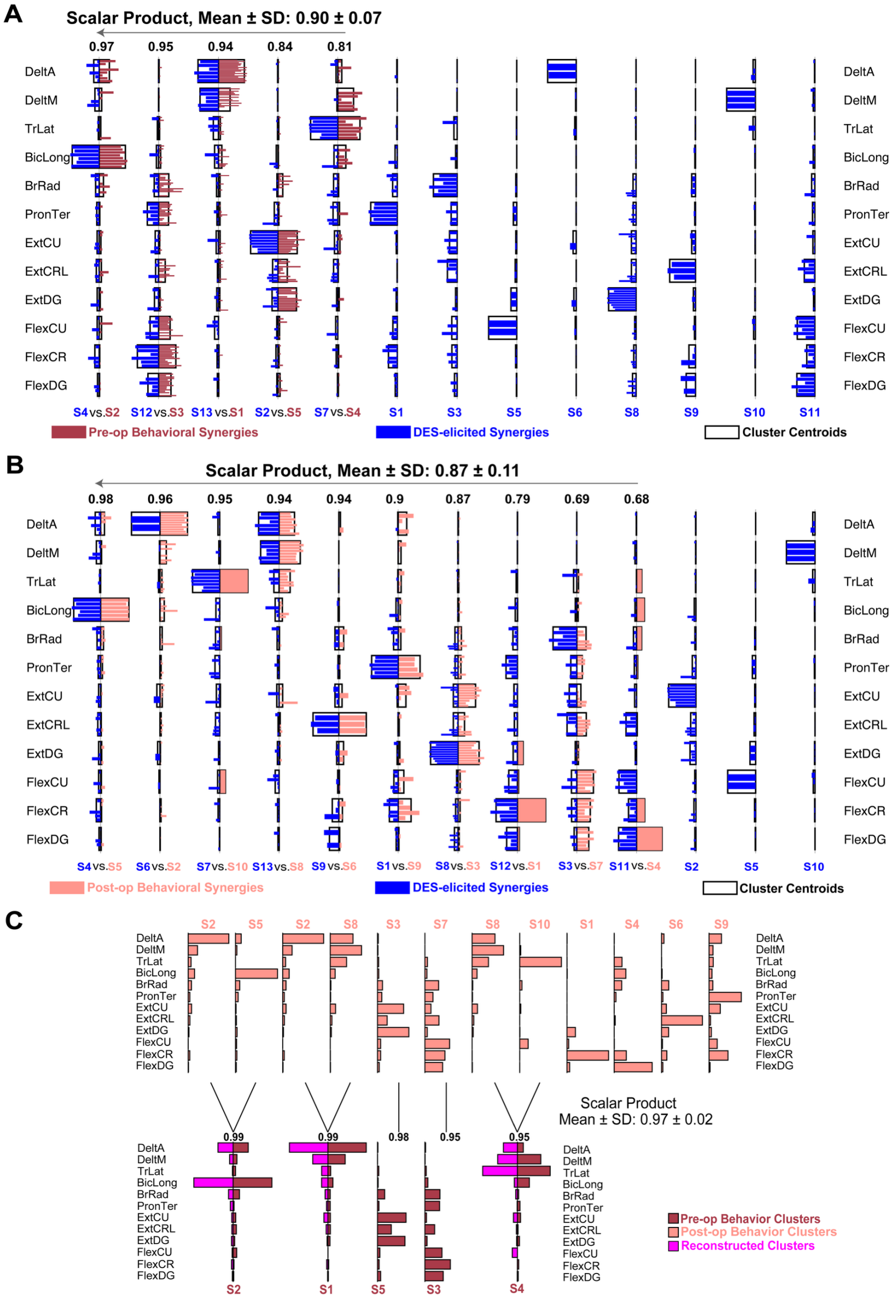


**Supplementary Figure 1. The pre- and post-op muscle synergy clusters and the DES-evoked synergy clusters are comparable. (A)** We *k*-means-clustered the pre-op behavioral muscle synergies (dark pink) and DES-evoked muscle synergies (blue) of all subjects, respectively, and matched each pre-op behavioral cluster to a DES-evoked cluster. All pre-op synergies could be paired with a DES-evoked synergy with moderate-to-high scalar product similarity (0.90 ± 0.07, mean ± SD). In each cluster pair, individual muscle synergies are plotted as bars with filled colors while the cluster centroids are shown as unfilled bars. The scalar product value between the cluster centroids of each pair is shown above the pair. **(B)** Comparison of the post-op behavioral muscle synergy clusters (light pink) and the DES-evoked synergy clusters (blue). This comparison yielded results similar to those presented in (A), though the average scalar product similarity between the cluster centroids was lower (0.87 ± 0.11). **(C)** When comparing the pre- and post-op behavioral muscle synergy clusters, some of the pre-op cluster centroids could be better explained by linearly combining multiple post-op cluster centroids. For instance, the pre-op cluster S2 (dark pink) could be reconstructed by merging the post-op clusters S2 and S5 (light pink), so that the scalar product between the pre-op S2 and its reconstruction (magenta) was 0.99.

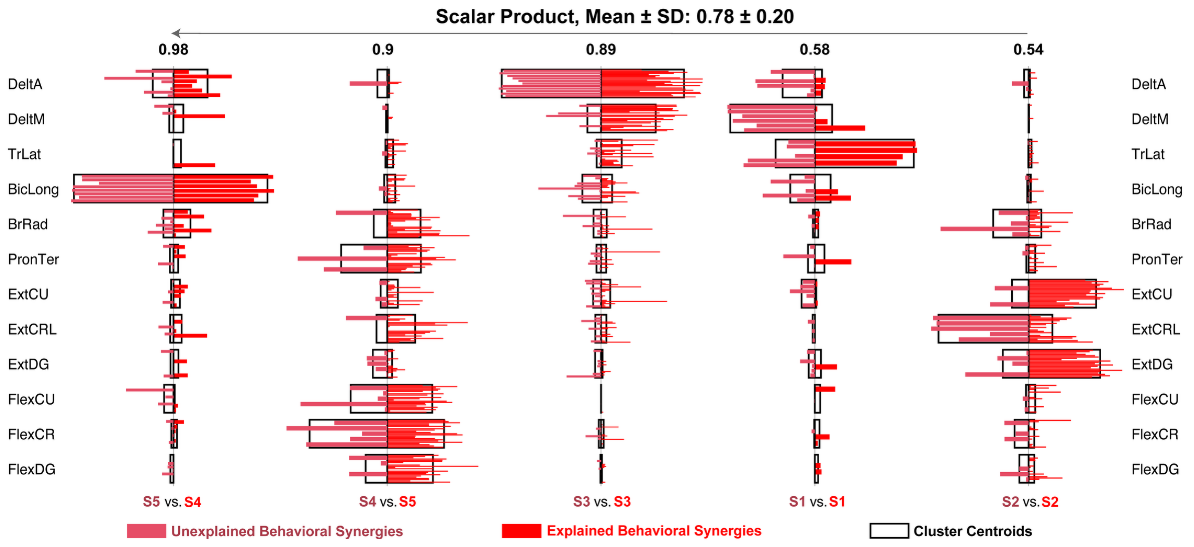


**Supplementary Figure 2. Comparison of the behavioral muscle synergies that were explained or unexplained by the DES-evoked synergies.** We *k*-means-clustered the explained pre- and post-op behavioral muscle synergies (red) and the unexplained behavioral muscle synergies (light red) of all subjects, respectively, and then compared the two groups of synergy clusters by assessing the similarity of the cluster centroids. The 5 pairs of synergy clusters showed a moderate-to-high scalar product (SP) similarity (0.78 ± 0.20, mean ± SD; range of 0.54 to 0.98), with a mean SP that was significantly higher than the baseline SP between cluster sets derived from shuffled explained and unexplained behavioral synergies (0.60 ± 0.12, mean ± SD; range of 0.48 to 0.79).In each cluster pair, individual muscle synergies are plotted as bars with filled colors while the cluster centroids are shown as unfilled bars. The scalar product value between the cluster centroids of each pair is shown above the pair.

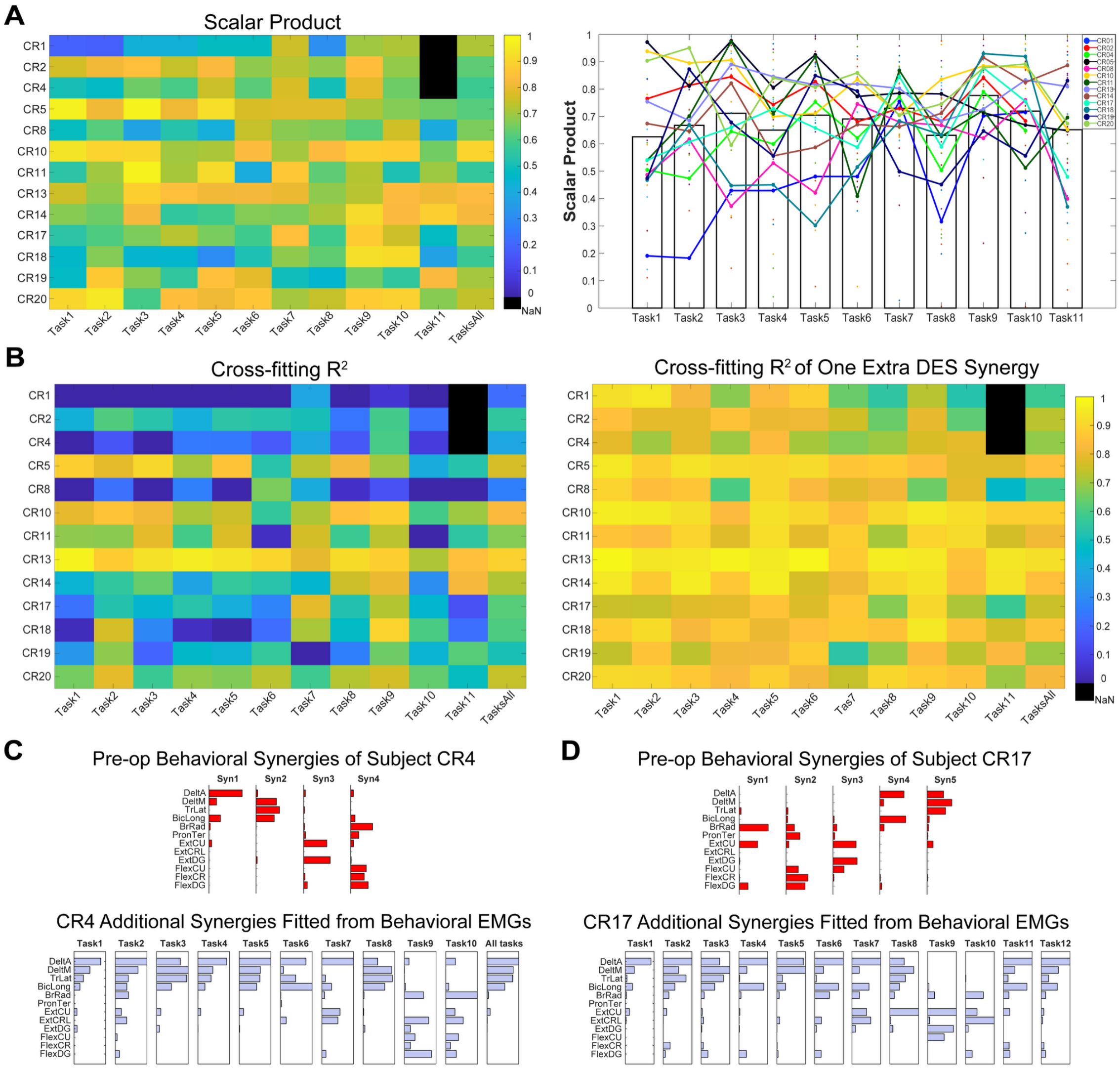


**Supplementary Figure 3. Similarity between the DES-evoked and pre-op behavioral muscle synergy sets across motor tasks**. **(A)** For each subject, similarity between the DES-evoked and pre-op behavioral muscle synergies were quantified using the scalar product (SP) between the best-matched DES-behavioral synergy pairs. Left panel, a heatmap showing the SP (averaged across all matched synergy pairs) for each subject, for each task and for all tasks combined (rightmost column). Entries with no data available are shown in black. Right panel, a bar plot that presents the SP, averaged across all subjects, for each behavioral task. Superimposed onto the bars are colored markers that represent the SPs of individual subjects; data points from each subject are joined together by straight lines of a unique color. **(B)** Similarity between the DES-evoked and pre-op behavioral synergy sets, evaluated by fitting the DES-evoked muscle synergy sets to the behavioral EMGs of each task. Left panel, the $R^2$ for the DES synergy-to-behavioral EMG cross-fitting for each subject, for each task, and for all tasks combined (rightmost column). Right panel, same as left panel, except that the cross-fit was performed by concurrently extracting one extra muscle synergy from the behavioral EMGs. As a result, the cross-fit $R^2$ became higher, as expected. **(C)** **(D)** Comparison of the pre-op behavioral muscle synergies and the additional muscle synergies extracted from the pre-op behavioral EMG as the DES-evoked synergies were fit to the data, for two subjects (CR4 in C, CR17 in D). It is apparent that for each task, the extra muscle synergy (light blue) resembles one of the behavioral muscle synergies (red). The extra synergies may therefore represent the behavioral synergies that were not retrieved by DES, probably due to the lack of an exhaustive coverage of M1 during motor mapping.

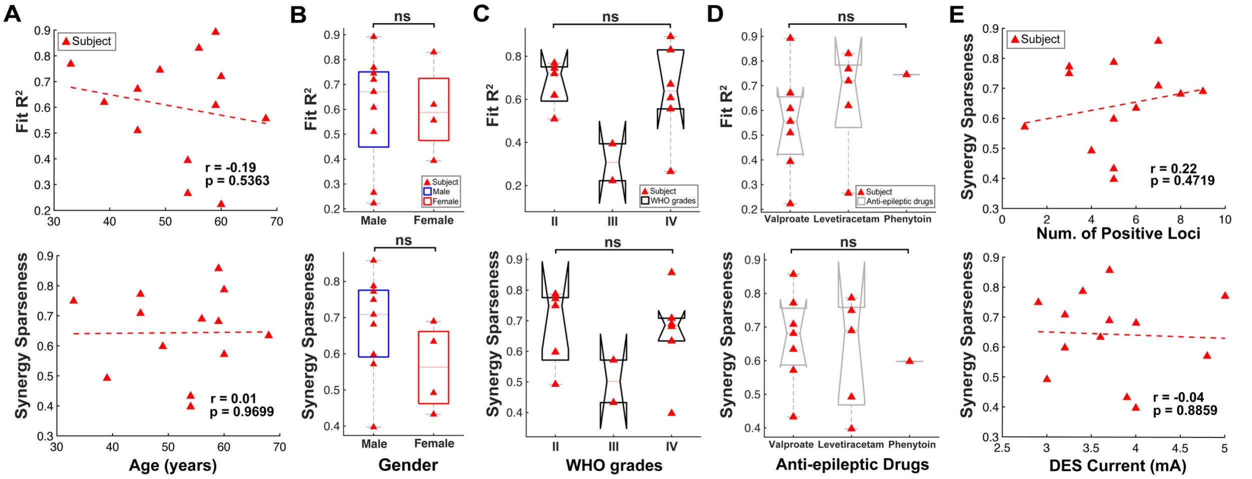


**Supplementary Figure 4. Demographic and experimental factors that might influence the similarity between the DES-evoked and behavioral muscle synergy sets. (A)** Across subjects, the subject's age did not correlate with either the DES synergy-to-behavioral EMG cross-fitting $R^2$ (top panel), or the average sparseness of the DES-evoked synergies (bottom panel). The Pearson's correlation coefficient (r) and its *p* value are shown on each correlation panel, with the dashed lines indicating insignificant relationships ($p > 0.05$). **(B)** Both the DES synergy-to-behavioral EMG cross-fitting $R^2$ (top panel) and the average sparseness of the DES-evoked synergies (bottom panel) did not differ significantly between males and females (ns, not significant, independent t-test). **(C)** WHO grades of brain tumor (II, III and IV) did not affect the DES synergy-to-behavioral EMG cross-fitting $R^2$ (top panel) or the average sparseness of the DES-evoked synergies (bottom panel) (ns, not significant; Kruskal-Wallis test). **(D)** Prescription of different anti-epileptic drugs (valproate, levetiracetam and phenytoin) did not influence the DES synergy-to-behavioral EMG cross-fitting $R^2$ (top panel) or the average sparseness of the DES-evoked synergies (bottom panel) (ns, not significant; Kruskal-Wallis test). **(E)** Neither the number of positive loci (top panel) nor the subject's stimulation current (bottom panel) correlated with the average sparseness of the DES-evoked muscle synergies.

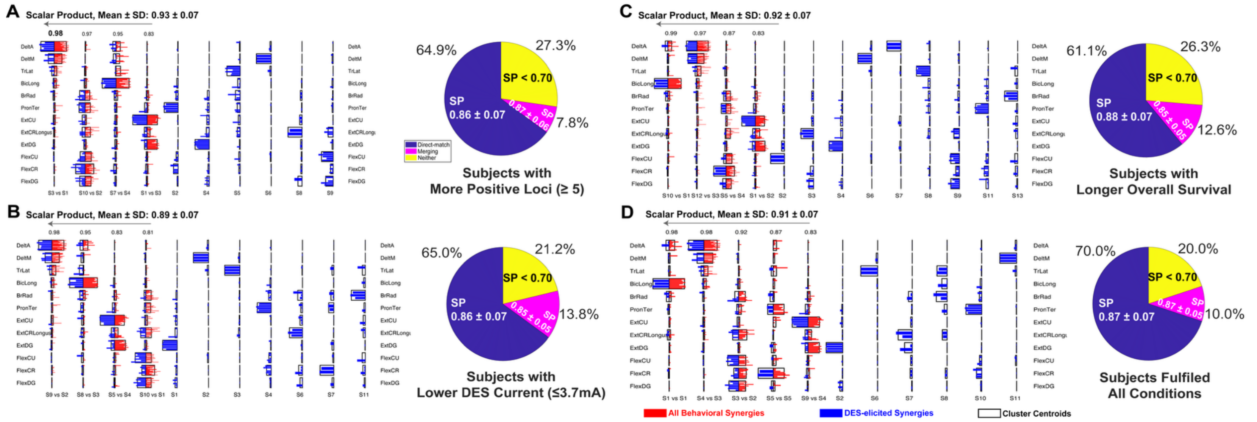


**Supplementary Figure 5. Similarity between behavioral and DES-evoked muscle synergies in select subgroups of subjects.** Subgroups of subjects having certain demographic and/or experimental characteristics were chosen for additional synergy analysis to gain insights on what factors may influence DES-behavioral muscle synergy similarity. In each subgroup, the DES-evoked and pre-op behavioral muscle synergies of the subjects were *k*-means-clustered separately. The two groups of clusters were then matched as synergy pairs based on the scalar product similarity between the cluster centroids. In each plot, individual muscle synergies are shown as colored bars (blue, DES-evoked synergies; red, behavioral synergies), and the cluster centroids are shown as unfilled bars superimposed onto the colored bars. The scalar product value of each synergy pair is shown on top of the pair. To the right of each synergy plot is a pie chart that shows the percentages of individual behavioral muscle synergies within the subgroup that (i) could be well matched (scalar product ≥ 0.7) directly to a DES-evoked synergy of the same subject (blue), (ii) could only be accounted for by merging multiple DES-evoked synergies of the same subject (magenta), and (iii) could not be explained (scalar product < 0.7) by either of the above (yellow). **(A)** Subject subgroup with more positive loci (≥5, n = 9). **(B)** Subject subgroup with lower stimulation current used (≤3.7 mA, n = 8). **(C)** Subject subgroup with longer post-op overall survival (i.e., with post-op assessment performed, n = 9). **(D)** Subject subgroup with more positive loci, lower stimulation current, and longer survival (n = 6). Note that with the above subject selection criteria imposed, the percentage of behavioral synergies that could not be explained by either direct matching or merging (i.e., the "neither" portion in yellow in the pie chart) decreased (cf. "neither" = 31% in Fig. 2D).

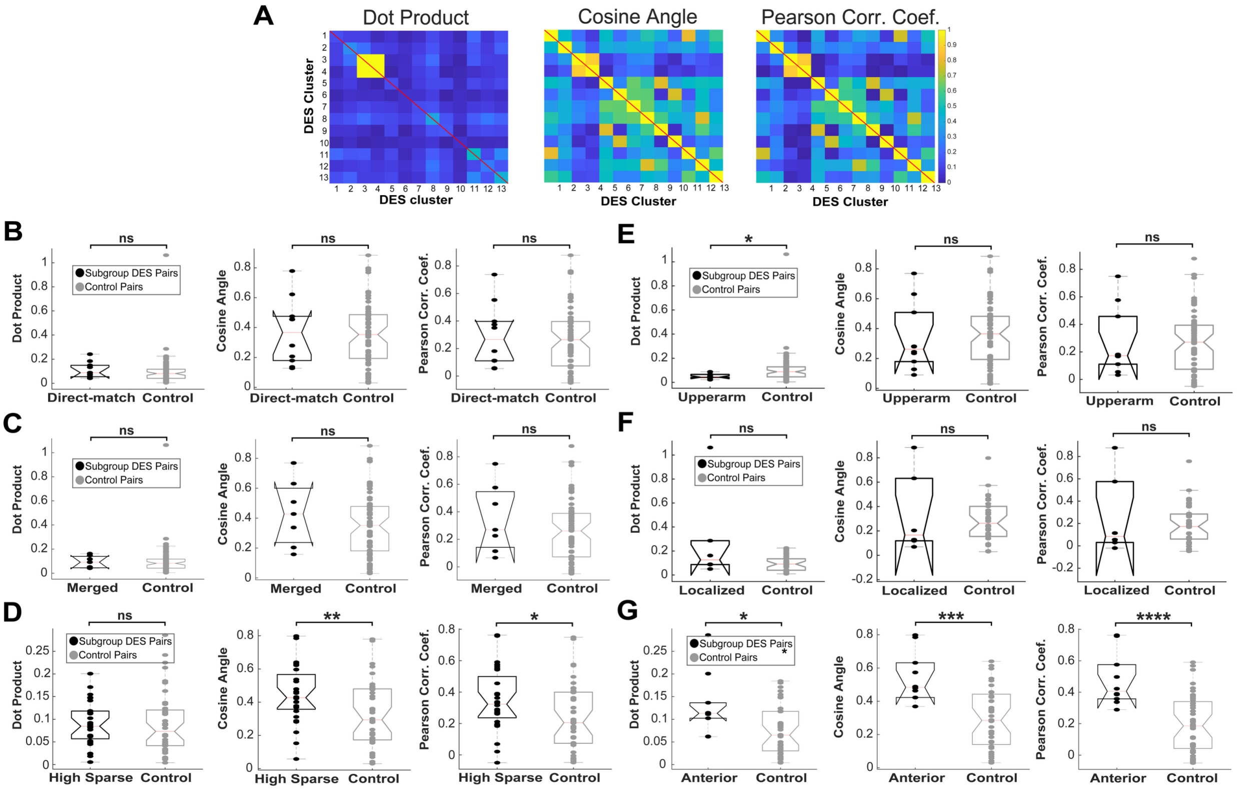


**Supplementary Figure 6. Similarity between the cortical activity maps of the different DES-evoked muscle synergy clusters. (A)** Similarity between any two of the cortical activity maps of the 13 DES-evoked muscle synergy clusters were evaluated by three measures: dot product (left panel), cosine angle (middle panel), and the Pearson correlation coefficient (right panel). For each measure, similarity values for all pairs of maps are shown as a heatmap. Note the variability of similarity values present in all measures. **(B)** Similarity between any two cortical activity maps of the DES-evoked muscle synergy clusters that could be well-matched directly to a behavioral synergy cluster ("Direct-match"), vs. the pairwise similarity of all other pairs of cortical activity maps (i.e., including pairs of unmatched clusters, and a matched plus an unmatched cluster) ("Control"). Comparisons using the three measures are presented (dot product, left panel; cosine angle, middle; Pearson correlation coefficient, right). **(C)** Pairwise similarity between cortical activity maps of DES-evoked synergy clusters that could be merged with another DES-evoked cluster for explaining a behavioral synergy cluster ("Merged"), vs. the pairwise similarity of all other pairs of cortical activity maps ("Control"), evaluated with the same three measures. **(D)** Pairwise cortical map similarity for pairs of clusters with high synergy sparseness ("High Sparse"), vs. all other pairs ("Control"). The cortical maps of the high-sparseness muscle synergies tended to be more similar to each other (**, $p < 0.01$; *, $p < 0.05$; independent t-test). **(E)** Pairwise cortical map similarity for pairs of clusters that involved upper arm muscles ("Upperarm"), vs. all other pairs ("Control"). Note that in the panel for dot product, the mean for the upper-arm synergies is lower than the control value (*, $p < 0.05$, independent t-test). **(F)** Pairwise cortical map similarity for pairs of clusters with localized cortical distributions ("Localized"), vs. all other pairs ("Control"). **(G)** Pairwise cortical map similarity for pairs of synergy clusters with an anterior activity distribution, as indicated by an anterior CoM position ("Anterior"), vs. all other pairs of clusters (“Control”). For all three measures, the muscle synergies with an anterior activity distribution were more similar to each other than to the rest (*, $p < 0.05$; ***, $p < 0.001$; ****, $p < 0.0001$; independent t-test). Abbreviation: ns, not significant.
